# Immuno-epigenetic signature derived in saliva associates with the encephalopathy of prematurity and perinatal inflammatory disorders

**DOI:** 10.1101/2022.10.18.22281194

**Authors:** Eleanor L.S. Conole, Kadi Vaher, Manuel Blesa Cabez, Gemma Sullivan, Anna J. Stevenson, Jill Hall, Lee Murphy, Michael J. Thrippleton, Alan J. Quigley, Mark E. Bastin, Veronique E. Miron, Heather C. Whalley, Riccardo E. Marioni, James P. Boardman, Simon R. Cox

**Affiliations:** Lothian Birth Cohorts group, Department of Psychology, University of Edinburgh, Edinburgh EH8 9JZ, UK; Centre for Genomic and Experimental Medicine, Institute of Genetics and Cancer, University of Edinburgh, Edinburgh EH4 2XU, UK; Centre for Clinical Brain Sciences, University of Edinburgh, Edinburgh EH16 4SB, UK; MRC Centre for Reproductive Health, Queen’s Medical Research Institute, Edinburgh BioQuarter, University of Edinburgh, Edinburgh, EH16 4TJ; Edinburgh Clinical Research Facility, University of Edinburgh, Edinburgh EH4 2XU, UK; Imaging Department, Royal Hospital for Children and Young People, Edinburgh, EH16 4TJ

**Keywords:** Perinatal, Inflammation, DNA methylation, Epigenetics, Encephalopathy of Prematurity, White Matter, Preterm Birth, Necrotizing Enterocolitis, Neonatal Sepsis, Diffusion Tensor Imaging, Neurodevelopment, Multiomics

## Abstract

**Background:** Preterm birth is closely associated with a phenotype that includes brain dysmaturation and neurocognitive impairment, commonly termed Encephalopathy of Prematurity (EoP), of which systemic inflammation is considered a key driver. DNA methylation (DNAm) signatures of inflammation from peripheral blood associate with poor brain imaging outcomes in adult cohorts. However, the robustness of DNAm inflammatory scores in infancy, their relation to comorbidities of preterm birth characterised by inflammation, neonatal neuroimaging metrics of EoP, and saliva cross-tissue applicability are unknown.

**Methods:** Using salivary DNAm from 258 neonates (n = 155 preterm, gestational age at birth 23.28 – 34.84 weeks, n = 103 term, gestational age at birth 37.00 – 42.14 weeks), we investigated the impact of a DNAm surrogate for C-reactive protein (DNAm CRP) on brain structure and other clinically defined inflammatory exposures. We assessed i) if DNAm CRP estimates varied between preterm infants at term equivalent age and term infants, ii) how DNAm CRP related to different types of inflammatory exposure (maternal, fetal and postnatal) and iii) whether elevated DNAm CRP associated with poorer measures of neonatal brain volume and white matter connectivity.

**Results:** Higher DNAm CRP was linked to preterm status (−0.0107 ± 0.0008, compared with - 0.0118 ± 0.0006 among term infants; p < 0.001), as well as perinatal inflammatory diseases, including histologic chorioamnionitis, sepsis, bronchopulmonary dysplasia, and necrotising enterocolitis (OR range |2.00 | to |4.71|, p < 0.01). Preterm infants with higher DNAm CRP scores had lower brain volume in deep grey matter, white matter, and hippocampi and amygdalae (β range |0.185| to |0.218|). No such associations were observed for term infants. Association magnitudes were largest for measures of white matter microstructure among preterms, where elevated epigenetic inflammation associated with poorer global measures of white matter integrity (β range |0.206| to |0.371|), independent of other confounding exposures.

**Conclusions:** Epigenetic biomarkers of inflammation provide an index of innate immunity in relation to neonatal health. Such DNAm measures complement biological and clinical metrics when investigating the determinants of neurodevelopmental differences.

## 1. Introduction

Preterm infants are at an increased risk of elevated inflammation, related health complications, and adverse neurodevelopment compared to infants born at term (1–8). While the aetiology of these outcomes is multifactorial, inflammation is considered to be a key component linking preterm birth and poor neurodevelopmental and mental health outcomes via its effects on cerebral maturational processes (9–12). Neonatal neuroimaging has identified neurostructural hallmarks of preterm birth commonly referred to as Encephalopathy of Prematurity (EoP), including dysmaturation of cortical and deep grey matter, atypical white matter development and disrupted connectivity (13). Recent advances in epigenetics may permit new ways to characterise sustained inflammation and reveal new insights into the relationship between inflammatory exposures, inflammation and neonatal brain and health outcomes.

Preterm infants are more susceptible to sustained inflammation than term infants and can be subject to multiple inflammatory stimuli during the perinatal period (14,15). Alongside maternal lifestyle-related exposures (16), various complications during pregnancy such as preeclampsia and histologic chorioamnionitis (17,18) can induce both maternal and fetal inflammatory responses and increase the risk of a sustained pro-inflammatory state postnatally (17–22). Preterm infants are additionally at higher risk for developing severe inflammatory conditions in the first few weeks of life, which may in turn perpetuate inflammation (8) – including bronchopulmonary dysplasia (BPD), necrotizing enterocolitis (NEC), severe retinopathy of prematurity (ROP) and episodes of sepsis (23). Preterm infants often present with multiple chronic conditions at once (24), putting them at higher risk of a greater allostatic load of inflammation (14,24–26).

Though numerous studies report associations between inflammation and cognitive outcomes in preterm populations (8,27–31), research linking inflammatory biomarkers with neurostructural measures yield inconsistent findings (4,6,11,19,21,32–37). Gaining a clearer understanding of the pathways via which sustained inflammation in very early life may precipitate well characterised cognitive and neurostructural outcomes requires novel approaches. The relative inconsistency of work to date is likely due to heterogeneity in study design: there is substantial variation in the demographic characteristics of study samples; the degree to which residual confounding factors are controlled; and the presence or absence of term control groups. Moreover, the relative nascency of the neonatal neuroimaging field (38) contributes to substantial variation in the acquisition and selection of brain outcome measures, and there is marked anatomic variation in early life, which can confound investigation of structure-function relationships (39).

Above these factors, we argue that the measures used to characterise inflammation in the first place may account for the greatest source of ambiguity in the inflammation-brain structure literature. In both clinical and research settings, there is a historical reliance on sampling phasic inflammation-related protein measures from blood to signpost inflammation. Of these, C-Reactive Protein (CRP) is the most widely adopted (40), although there are criticisms of this approach (41,42), particularly in newborn populations (40,43) where the CRP response to inflammation is variable owing to an immature immune system (15). Accurate characterisation of sustained (and not transient or acute) inflammation arguably requires repeated sampling or follow up examinations, which few studies endeavour to profile (44). This, alongside uncertainty of what exactly constitutes sustained inflammation in the preterm infant (15) calls for alternative, more stable biomarkers which can accurately reflect baseline inflammation levels. There is a clear precedent to identify tools to both circumvent the practical limitations of conventional inflammatory measurement in the neonate, alongside study designs that necessitate detailed brain and clinical information with adequate statistical power to detect small to moderate effect sizes.

Our previous work demonstrated that DNA methylation (DNAm) markers of inflammation may provide more stable readouts of cumulative inflammatory exposure (45,46) and shed greater insight into the consequences of inflammation on brain structure (47,48). DNAm is an epigenetic mechanism that can act as an interface by which environmental exposures influence gene function. DNAm is dynamic during fetal development, both in terms of the developing immune system (49) and brain (50–52) and may mediate the impact of maternal, fetal and postnatal exposures on brain development (53–55). In the context of preterm birth, only a limited number of studies have investigated DNAm changes (53,56–65) – of these, few examine DNAm in relation to neonatal neuroimaging metrics (59,62,63). Additionally, though some of these studies have examined DNAm in relation to postnatal health outcomes (53,65), no study to date has examined inflammation, DNAm, and neuroimaging concurrently in the neonatal period. This study is the first time an epigenetic measure of inflammation has been examined in a neonatal cohort in relation to brain health outcomes.

Here, using a cohort of 258 infants (103 term, 155 preterm), we examine (1) how a salivary DNAm signature of the inflammatory marker C-reactive protein (DNAm CRP) relates to preterm birth (2) how this signature associates with maternal, fetal and postnatal inflammatory exposures both individually and in aggregate, and (3) how variance in this measure relates to global measures of MRI brain volume, diffusion MRI (dMRI) correlates of connectivity, and regional variation in individual white matter tracts.

## 2. Methods

### 2.1 Study population

Preterm (gestational age at birth < 37 weeks) and term born infants delivered at the Royal Infirmary of Edinburgh, UK were recruited to the Theirworld Edinburgh Birth Cohort, a longitudinal study designed to investigate the effect of preterm birth on brain development (66). Cohort exclusion criteria were major congenital malformations, chromosomal abnormalities, congenital infection, overt parenchymal lesions (cystic periventricular leukomalacia, hemorrhagic parenchymal infarction) or post-hemorrhagic ventricular dilatation. Ethical approval has been obtained from the National Research Ethics Service, South East Scotland Research Ethics Committee (11/55/0061, 13/SS/0143 and 16/SS/0154). Informed consent was obtained from a person with parental responsibility for each participant. DNAm data were available from 258 neonates, 214 of whom also had successful structural and diffusion MRI acquisition.

### 2.2 Study variables

Inflammatory exposures were coded as binary variables (1 = present, 0 = absent) and were grouped as follows: maternal (pertaining to mother / maternal exposure), fetal (affecting placenta or fetus) or neonatal (affecting infant after birth). **Table 1** presents participant characteristics of these categories. Histologic chorioamnionitis (HCA) was defined via placental histopathology, as reported previously (17,21). Incidence of any neonatal sepsis (either late onset or early onset sepsis) was defined as detection of bacterial pathogen from blood culture, or physician decision to treat for ≥5 days in the context of growth of coagulase negative staphylococcus from blood or a negative culture. Necrotising enterocolitis (NEC) was defined as stages two or three according to the modified Bell’s staging for NEC (67). Bronchopulmonary dysplasia (BPD) was defined by the requirement for supplemental oxygen at 36 weeks gestational age. Birthweight z-scores were calculated according to International Fetal and Newborn Growth Consortium for the 21st Century (INTERGROWTH-21st) standards (68).

**Table 1.** Demographic and clinical features of study sample (n=258). P values denote significant difference between term and preterm groups.

| Characteristics & clinical features | Term infants (n=103) | Preterm infants (n=155) | P value |
| --- | --- | --- | --- |
| Sex: Female (%) | 44 (43) | 75 (48) | 0.2166 |
| Gestational age at birth/weeks (range) | 39.7 (37.00 – 42.14) | 28.84 (23.28 – 34.84) | <0.001 |
| Gestational age at scan/weeks (range) | 42.27 (39.84 – 47.14) | 40.56 (37.70 – 45.14) | <0.001 |
| Birth weight/g (range) | 3482 (2346 – 4670) | 1177 (500 – 2100) | <0.001 |
| Birth weight z-score (range) | 0.43 (-2.30 – 2.96) | -0.19 (-3.13 – 1.58) | <0.001 |
| DNAm CRP (mean, SD) | -0.012 (0.001) | -0.011 (0.001) | <0.001 |
| **Maternal / fetal** |  |  |  |
| Maternal age (years) | 33.7 (19–48) | 31.1 (17–44) | <0.001 |
| Antenatal corticosteroid administration in pregnancy, n (%) | 2 (0.02) | 148 (95.5) | <0.001 |
| MgSO4 administration in pregnancy, n (%) | 0 (0) | 112 (72) | <0.001 |
| Smoked during pregnancy (%) | 2 (2) | 29 (19) | <0.001 |
| Histologic chorioamnionitis, n (%) | 6 (0.06) | 49 (31.6) | <0.001 |
| Preeclampsia, n (%) | 7 (0.07) | 22 (14.2) | <0.001 |
| **Neonatal** |  |  |  |
| Necrotizing enterocolitis, n (%) | 0 (0) | 10 (6) | N/A |
| Bronchopulmonary dysplasia, n (%) | 0 (0) | 48 (31) | N/A |
| Retinopathy of prematurity, n (%) | 0 (0) | 8 (5) | N/A |
| Sepsis, n (%) | 0 (0) | 36 (23) | N/A |

### 2.3 DNA extraction and methylation measurement and pre-processing

Saliva obtained at term equivalent age was collected in Oragene OG-575 Assisted Collection kits, by DNA Genotek, and DNA extracted using prepIT.L2P reagent (DNA Genotek, Ontario, Canada). DNA was bisulfite converted and methylation levels were measured using Illumina HumanMethylationEPIC BeadChip (Illumina, San Diego, CA, USA) at the Edinburgh Clinical Research Facility (Edinburgh, UK). The arrays were imaged on the Illumina iScan or HiScan platform and genotypes were called automatically using GenomeStudio Analysis software version 2011.1 (Illumina). Infants’ saliva samples were taken (for DNAm analysis) around the same time as MRI acquisition. Details of DNAm pre-processing have been outlined previously (63); for full details, refer to **supplementary methods**.

### 2.4 Inflammatory-related methylation signature

For each individual (n = 258), a weighted linear signature (DNAm CRP) was obtained by multiplying the methylation proportion at a given CpG by the effect size from a previous epigenome wide association study (EWAS) of CRP (69) (**supplementary table 1**), and then summing these values. This method has been described previously (46–48,70).

### 2.5 MRI acquisition

This study incorporates data from two phases of MRI acquisition which is reflected in the flowchart of the study sample **(supplementary figure 2)**. The data acquisition of this study has been reported previously (63,66).

In the first phase (n = 93), structural and dMRI were performed in neonates using a MAGNETOM Verio 3T clinical MRI scanner (Siemens Healthcare GmbH, Erlangen, Germany) and 12-channel phased-array head coil. For dMRI, A protocol consisting of 11 baseline volumes (b = 0 s/mm^2^ [b0]) and 64 diffusion-weighted (b = 750 s/mm^2^) single-shot spin-echo echo planar imaging (EPI) volumes acquired with 2 mm isotropic voxels (TR/TE 7300/106 ms) was used; 3D T1-weighted (T1w) MPRAGE (TR/TE 1650/2.43 ms) with 1 mm isotropic voxels was acquired.

For the second phase (n=121), structural and dMRI were performed neonates using a MAGNETOM Prisma 3T clinical MRI scanner (Siemens Healthcare GmbH, Erlangen, Germany) and 16-channel phased-array pediatric head and neck coil. This was used to acquire dMRI in two separate acquisitions: the first consisted of 8 b0 and 64 volumes with b = 750s/mm^2^; the second consisted of 8 b0, 3 volumes with b = 200 s/mm^2^, 6 volumes with b = 500s/mm^2^ and 64 volumes with b = 2500 s/mm^2^. An optimal angular coverage for the sampling scheme was applied (71). In addition, an acquisition of 3 b0 volumes with an inverse phase encoding direction was performed. All dMRI volumes were acquired using single-shot spin-echo planar imaging (EPI) with 2-fold simultaneous multi-slice and 2-fold in-plane parallel imaging acceleration and 2 mm isotropic voxels; all three diffusion acquisitions had the same parameters (TR/TE 3500/78.0ms). Images affected by motion artifact were re-acquired multiple times as required; dMRI acquisitions were repeated if signal loss was seen in 3 or more volumes. 3D T2-weighted SPACE images (T2w) (TR/TE 3200/409 ms) with 1 mm isotropic voxels and 3D T1w MPRAGE (TR/TE 1970/4.69 ms) with 1 mm isotropic voxels were also acquired.

Infants were fed and wrapped and allowed to sleep naturally in the scanner without sedation. Pulse oximetry, electrocardiography and temperature were monitored. Flexible earplugs and neonatal earmuffs (MiniMuffs, Natus) were used for acoustic protection. All scans were supervised by a doctor or nurse trained in neonatal resuscitation. Structural images were reported by an experienced pediatric radiologist (A.J.Q), and each acquisition was inspected contemporaneously for motion artefact and repeated if there had been movement while the baby was still sleeping; dMRI acquisitions were repeated if signal loss was seen in 3 or more volumes.

As details on dMRI pre-processing have been previously outlined (72) please refer to **supplementary methods** for specifics. T2w images from phase 2 were processed using the dHCP pipeline (73). The T1w images from phase 1 were processed using specific software for brain skull-stripping and tissue segmentation (74). The phase 1 pipeline relies on some atlases, for these purposes, 10 subjects from the phase 2 that have both T1w and T2w were selected. The volumes extracted include cortical grey matter, deep grey matter, white matter, hippocampi and amygdalae, cerebellum, brainstem, cerebrospinal fluid (CSF) and ventricles.

From the diffusion images we calculated the tensor – fractional anisotropy (FA), mean diffusivity (MD), axial diffusivity (AD), and radial diffusivity (RD) – and the NODDI (intracellular volume fraction [NDI] maps) (75,76). All the subjects were registered to the Edinburgh Neonatal Atlas (ENA50) using DTI-TK (75,77). The diffusion tensor derived maps of each subject (FA and MD) were calculated after registration; NDI was then propagated to the template space using the previously calculated transformations. The data was skeletonized using the ENA50 skeleton and then multiplied by a custom mask. Finally, the peak width of the histogram of values computed within the skeletonized maps was calculated as the difference between the 95th and 5th percentiles (78). Global values of white matter microstructure reported in this study are the peak width of skeletonised metrics (PSFA, PSMD, PSRD, PSAD, PSNDI), which have been derived from the same pipeline previously used to characterise brain structural differences between preterm and term infants (77).

### 2.6 Tract segmentation and extraction of tract-averaged dMRI metrics

As above, details of individual white matter tract segmentation and subsequent extract of tract-averaged dMRI metrics have been outlined previously from infants from this study sample (79). Briefly, FA and MD were derived for the left and right hemispheric tracts of the arcuate fasciculus (AF), anterior thalamic radiation (ATR), cingulum cingulate gyrus (CCG), corticospinal tracts (CST), inferior fronto-occipital fasciculus (IFOF), inferior longitudinal fasciculus (ILF), uncinate fasciculus (UNC) and genu and splenium of the corpus callosum (CC).

### 2.7 Statistical Analysis

All statistical analyses were performed in R (version 4.0.5) (R Core Team, 2020).

#### 2.7.1 Selection of covariates and confounding variables

In comparing participant demographics between term infants (n = 103) and preterm infants (n =155), p-values derived from the t-test (for continuous variables) and Chi-square test or Fischer’s Exact test if the count was below 5 (categorical variables) (**Table 1**) (80). In all models examining the association between DNAm CRP with health outcomes and brain MRI metrics, we adjusted for infant sex, gestational age at birth, gestational age at scan, and birthweight Z score. Scanner variable was included when data included two phases of MRI acquisition. We also tested for an interaction between gestational age × DNAm CRP and infant sex × DNAm CRP in sensitivity analyses, where a significant interaction would indicate differences in association magnitudes at different gestational ages / between males and females. To quantify the amount of variance in each brain imaging biomarker accounted for by DNAm CRP, for all neuroimaging associations we report both the adjusted R^2^ and the incremental R^2^ (81), the latter of which was calculated by comparing the R^2^ of each model with that from a baseline model (reported as **Model H**_**0**_) R^2^ in which the MRI measure was modelled with covariates only (e.g., *Model H*_*0*_ *= MRI metric ∼ sex + birthweight + gestational age age + gestational age at scan + scanner*).

After examining global associations, we wanted to control for variables that could either cause the raised DNAm CRP (the exposure), variance in MRI metrics (the outcome), or both. We ran bivariate correlations and from these we included all common correlates of the exposure and the outcome in our second regression model (Model H_2_) to eliminate alternative explanations of the outcome due to confounding (**supplementary figure 1**). In these models, neither antenatal treatment of corticosteroids and MgSO4 for anticipated preterm birth were included to circumvent issues of multicollinearity, since they were given to the majority of mothers (96% and 72% respectively) in the preterm group and were highly correlated with preterm status (r = 0.71 – 0.93, p<0.001). The magnitude of effects are classified as small, medium, or large when the standardized coefficients are 0.1, 0.3, or 0.5, respectively as classified by Cohen (82).

Finally, accounting for the fact that inflammatory risk factors are positively correlated, we included all inflammatory risk factors in one multiple linear regression alongside DNAm CRP signature for each MRI variable of interest. This allowed us to account for unique contribution of DNAm CRP in the context of inflammatory-related exposures to variance in brain structural outcomes.

#### 2.7.2 Multiple inflammatory hits and DNAm CRP

We next performed investigations to assess whether DNAm was related to number of inflammatory episodes experienced. Due to small numbers of individual inflammatory risk factors and the frequent overlap of episodes experienced in the preterm group, we created binary outcome measures based on combinations of inflammatory risk factors or conditions experienced, combining infants that experienced three or more morbidities into a single group, resulting in four possible levels for the risk score of 0, 1, 2, or 3 + alongside a term control reference (0 inflammatory episodes). Results are presented firstly unadjusted (model H_1_), then adjusted for gestational age at birth, infant sex and birthweight z-score (model H_2_), and then adjusted for gestational age at birth, infant sex and birthweight z-score as well as administration of MgSO4 and corticosteroids in pregnancy (model H_3_), given these have been identified as potential confounders of the relationship between inflammation and health outcomes in previous studies (83–85). Results are presented as odds ratios (OR) and 95% confidence intervals (CI) for categorical outcome measures.

#### 2.7.3 DNAm CRP and global brain structure associations

To determine the effect of inflammation (DNAm CRP) on neuroimaging outcomes, data were analysed using regression models, controlling for gestational age at birth, gestational age at scan, infant sex, MRI scanner, and birthweight Z score. We aimed to contextualise these associations with clinical health data. Inflammatory risk factors were added simultaneously as covariates into a second model (model H_2_) in addition to the standard covariates of gestational age at birth, gestational age at scan, birthweight Z score and sex (model H_1_). As no term infants had postnatal inflammatory episodes (sepsis, NEC, ROP, BPD), analyses were stratified according to term or preterm status. In models testing global brain structural metrics such as brain volumes and PSMD and PSFA, MRI scanner was included as a binary covariate as MRI data from both phases of data collection were included (refer to **supplementary figure 2**, study sample flowchart). All continuous variables were standardised using z-score scaling to obtain standardised effect sizes (β). P-values were corrected for multiple testing using the false discovery rate (FDR) method and significance was deemed FDR corrected p-value (pFDR) < 0.05. 95% CIs are reported throughout.

#### 2.7.4 DNAm CRP and dMRI White matter tract associations

dMRI measures of white matter appear to be highly correlated (e.g. high FA in an individual tract such as the arcuate fasciculus is often accompanied by high FA across all other white matter tracts in that individual), a property that persists from early infancy through to older age (79,86,87). As a result of this, it is common to derive general factors (g-factors) of white matter microstructure to characterise global white matter microstructure. One PCA was conducted for FA and MD parameters across the 16 tracts to quantify the proportion of shared variance between them; in each analysis, each subject was described by 16 features, computed as the tract-averaged values of FA or MD across each tract (**supplementary figure 5**). The first unrotated principal component (PC) scores were extracted as the single-metric g-factors, gFA and gMD (scree plot and PCA variable contributions illustrated in **supplementary figure 6**).

As with global brain structural metrics, two models were used:

1. **Model H**_**1**_: dMRI metric ∼ DNAm CRP + gestational age at scan + gestational age at birth + infant sex + birthweight.
2. **Model H**_**2**_: dMRI metric ∼ DNAm CRP + gestational age at scan + gestational age at birth + infant sex + birthweight + all inflammatory risk factors

In comparison to global MRI volumetric metrics and PSFA, PSMD, PSAD and PSRD, all individual tract associations, gFA, gMD and PSNDI were limited to neuroimaging data from phase 2 of the study, hence no scanner variable was included in these analyses.

### 2.8 Data and code availability

Requests for original image and anonymised data will be considered through the BRAINS governance process (www.brainsimagebank.ac.uk). Raw DNAm data are available upon request from Theirworld Edinburgh Birth Cohort, University of Edinburgh (https://www.tebc.ed.ac.uk/2019/12/data-access-and-collaboration), while DNAm and metadata are not publicly available, generated DNAm CRP signatures are included alongside scripts for data analysis. All brain volumetric metrics were obtained using the scripts provided in https://github.com/amakropoulos/structural-pipeline-measures. The segmented tracts in the ENA50 template space are available online: https://git.ecdf.ed.ac.uk/jbrl/ena. Code for primary data analysis and figures are available at https://github.com/EleanorSC/TEBC_DNAmCRP and code for tract propagation and average calculation are available at https://git.ecdf.ed.ac.uk/jbrl/neonatal-gfactors.

## 3. Results

### 3.1 Participant characteristics

The study group consisted of 258 neonates: 155 participants were preterm and 103 were controls born at full term, see Table 1 for participant characteristics and **supplementary figure 1** for a flowchart of data acquisition. Among the preterm infants, 48 (31%) had bronchopulmonary dysplasia, 10 (6%) developed necrotising enterocolitis, 8 (5%) developed ROP, 49 had HCA (32%), 22 (15%) were born to women whose pregnancy was complicated by preeclampsia, and 36 (23%) had an episode of postnatal sepsis. Of the 258 participants with DNAm data, 214 also had MRI data.

### 3.2 Multiple inflammatory hits increase risk of elevated epigenetic inflammation

Preterm Infants in the sample for whom DNAm data and composite neonatal inflammatory risk scores were available (n = 155), had high prevalence (n = 112, 72%) of experiencing at least one of the documented inflammatory exposures (Figure 1B), which included incidence of smoking during pregnancy, preeclampsia, HCA, sepsis, BPD, NEC or ROP. A small subset of these infants experienced three or more of these exposures (n = 24, 15%).

**Figure 1.**
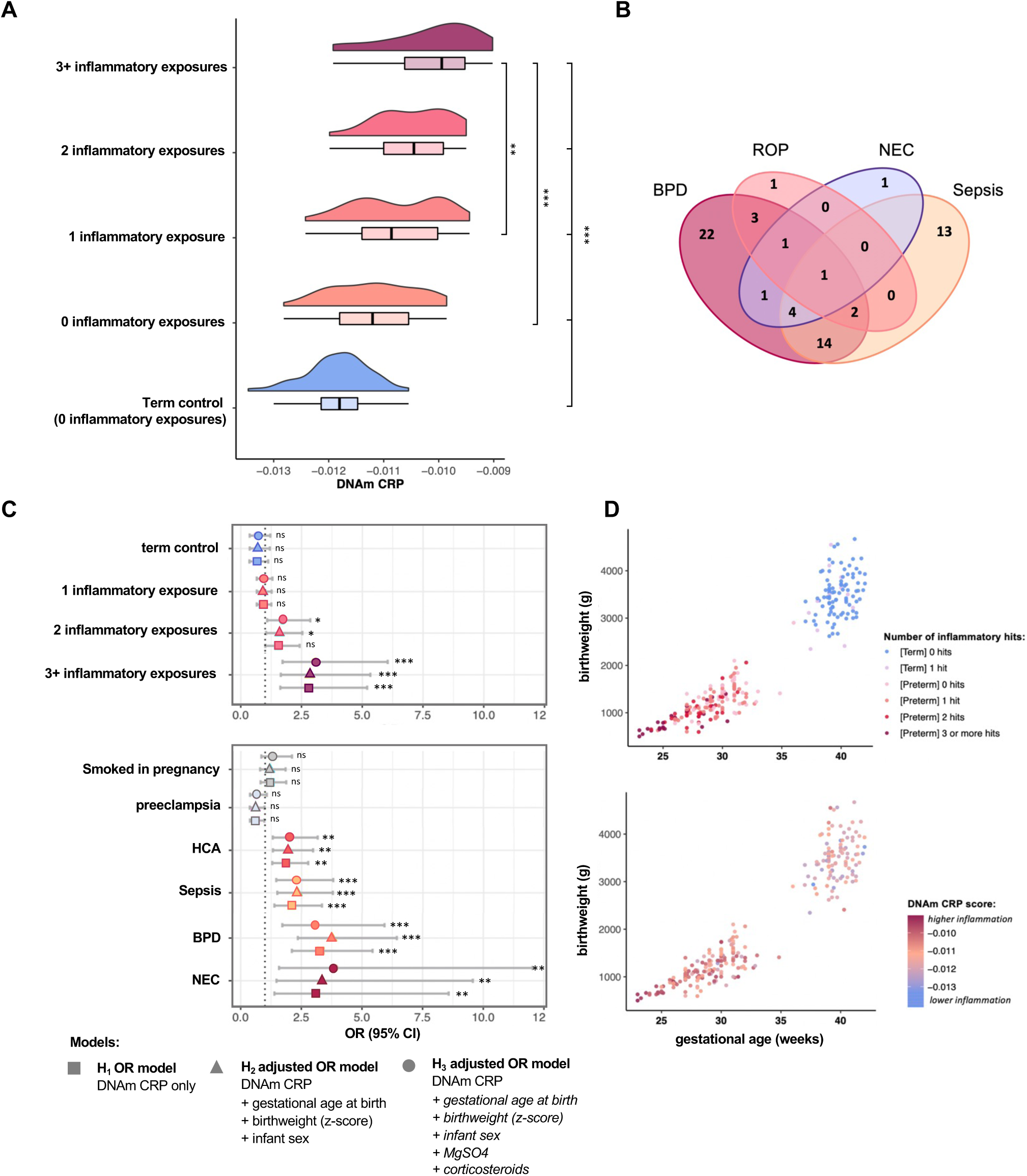
Multiple inflammatory hits associate with raised DNAm CRP. (A) distributions of DNAm CRP according to number of inflammatory episodes experienced by infant (B) Venn diagram showing the overlap postnatal inflammatory morbidities in study sample (C) Odds ratios and 95% confidence intervals for contribution of DNAm CRP to inflammatory exposures, asterisks (*) indicate statistically significant (FDR-corrected p < 0.05) (D) Scatter plots of the relationships between gestational age and birthweight, coloured according to number of inflammatory episodes/exposures (top panel) and DNAm CRP (bottom panel).

There was an association between number of inflammatory episodes and the epigenetic inflammation signature, with higher DNAm CRP in infants who had experienced greater exposure to inflammation. DNAm CRP was associated with higher odds of several perinatal morbidities including HCA, sepsis, BPD, and NEC. These relationships remained significant following adjustment for gestational age at birth, birthweight, and infant sex as well as perinatal variables of administration of corticosteroids and MgSO4 in pregnancy (Figure 1C, **supplementary table 3**). The association of DNAm CRP with ROP was no longer significant after controlling for MgSO4 and corticosteroid administration (model H_3_). DNAm CRP was also associated with three or more inflammatory episodes. There was no significant association of DNAm CRP with maternal smoking in pregnancy or preeclampsia. Infants with increasing numbers of complications were more likely to have shorter gestational ages at birth and lower birthweights (**Figure 1D**). Furthermore, preterm infants had significantly higher DNAm CRP (Table 1, p < 0.001). When examining DNAm CRP alongside clinical inflammatory exposures (**supplementary table 4**), there was no significant difference between term infants with no inflammatory episodes vs those with one. The largest difference was found between term infants with no inflammatory episodes and preterm infants with 3 or more inflammatory risk-factors (p < 0.001).

### 3.3 DNAm CRP and brain volumes

Overall, magnitudes of associations between DNAm CRP and global MRI brain volumes were modest, explaining a small amount of additional variance beyond covariates (of infant sex, gestational age at birth, birthweight z-score, gestational age at scan, scanner variable). After examining inflammation-brain structure associations across all infants (n = 214; **supplementary table 5**), we stratified analyses into term (n = 87) and preterm (n = 127) subgroups to examine group differences (Figure 2B). The incremental R^2^ upon adding DNAm CRP to a null model varied by trait: global white matter volume = 3.7%, deep grey matter volume = 3.0%, hippocampi and amygdalae volume = 2.7% and cerebellar volume = 1.6%. Visual inspection of diagnostic plots suggested no regression assumptions were violated (an example is given in **supplementary figure 4**). Term infants displayed no significant brain structural associations with DNAm CRP, whereas preterm infants with higher DNAm CRP displayed brain volume reductions in deep grey matter, white matter, and hippocampi and amygdalae (Figure 2B).

**Figure 2.**
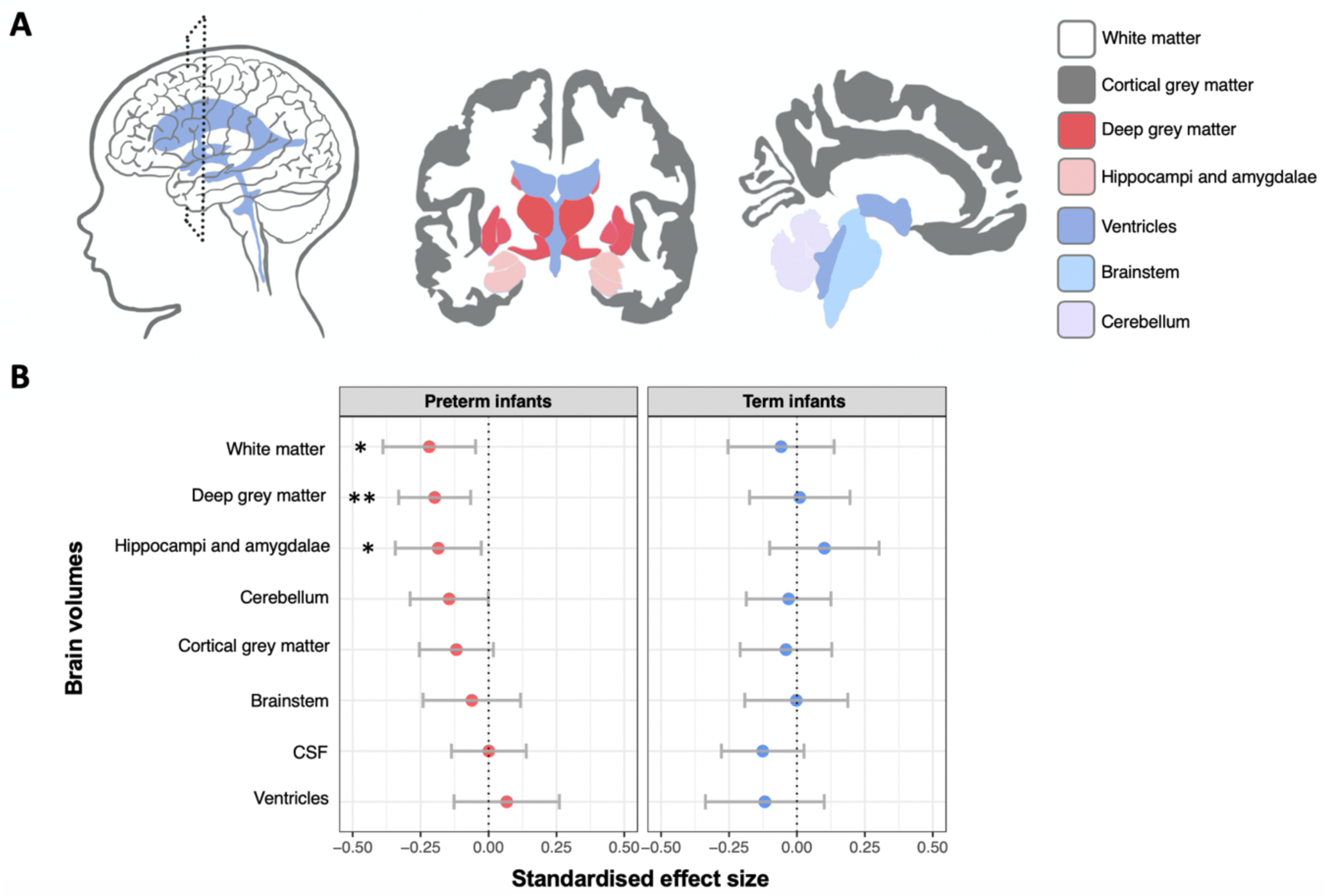
Association of DNAm CRP with brain volumes. (A) schematic drawn by ggseg package (Mowinckel & Vidal-Piñeiro, 2020) of reconstructed MRI brain volumetric measures (B) Standardized regression coefficients for DNAm CRP associations between brain volumetric measures for preterm infants (red circles) and preterm infants (blue circles). Points show standardized coefficients and 95% confidence intervals. Asterisks (*) indicate statistically significant (FDR-corrected p < 0.05). All models are controlled for infant sex, gestational age at birth, gestational age at scan and birthweight Z score and scanner variable.

Analyses were repeated to include interactions between DNAm CRP and both sex and gestational age (**supplementary tables 6-9**). While null findings were observed with the former (p > 0.05; **supplementary table 7**), within the preterm cohort there was evidence for interactions with gestational age at birth (**supplementary table 8**). Higher DNAm CRP was consistently associated with lower brain volume in infants of lower gestational ages (i.e. extremely preterm infants tended to have higher DNAm CRP and correspondingly smaller global brain volume measures). In contrast, there was no significant interaction between gestational age and DNAm CRP within the term sub-group (p > 0.05; **supplementary table 9**).

In fully adjusted models (Figure 3, **supplementary table 10**), there remained a significant association of DNAm CRP with deep grey matter volume (β = -0.209, p = 0.008), white matter volume (β = -0.304, p = 0.003), and cerebellum volume (β = -0.204, p = 0.013). For most brain metrics, the strength of the association between DNAm CRP and MRI metric was increased when additional inflammatory covariates were included in the model (percentage increase for deep grey matter volume = 6%, white matter volume = 39%, and cerebellum volume = 27%). Individually modelling risk factors revealed that this increase was mostly driven by controlling for incidence of sepsis, whereas brain structural associations were most attenuated by controlling for incidence of BPD (Figure 3).

**Figure 3.**
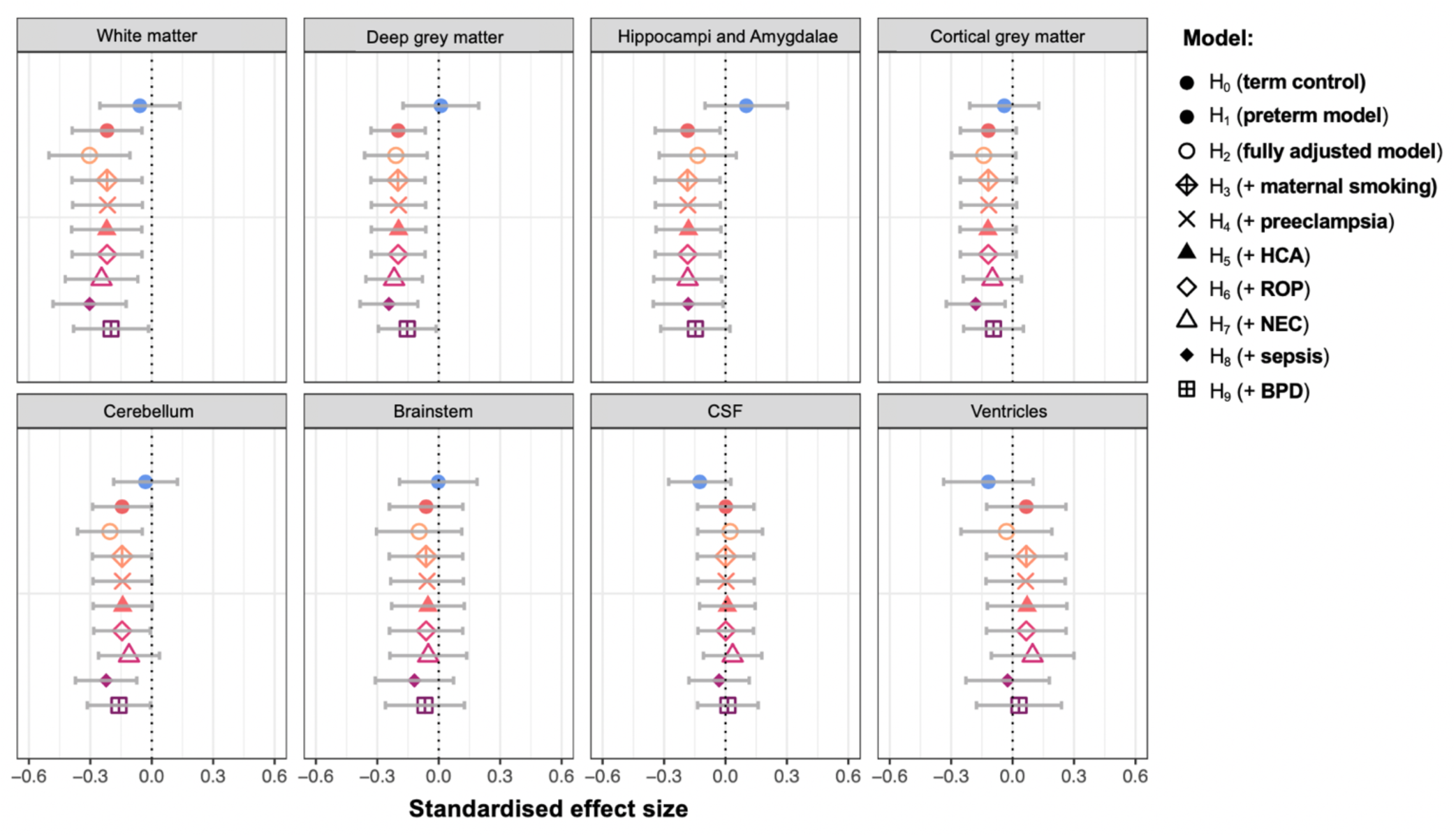
Associations between DNAm CRP and brain volumes and the impact of inflammatory risk factors on associations; Standardized regression coefficients for DNAm CRP associations between brain volumetric measures for preterm infants (red circles) and preterm infants (blue circles). Points show standardized coefficients and 95% confidence intervals. All models are controlled for infant sex, gestational age at birth, gestational age at scan and birthweight Z score and scanner variable; additional shapes show standardised regression coefficients for different models (H_2_-H_9_, corresponding with supplementary table 10).

### 3.4 Global white matter microstructure associations with DNAm CRP

Preterm infants with higher DNAm CRP had poorer measures of white matter tract integrity. This was seen at the global level for all peak width of skeletonised white matter microstructure metrics (**Table 2**); PSFA, PSMD, PSRD, PSAD; β range |0.186| to |0.341|, incremental R^2^ 2.7 – 9%). In term infants, there were no significant associations (**supplementary table 10**).

**Table 2.**
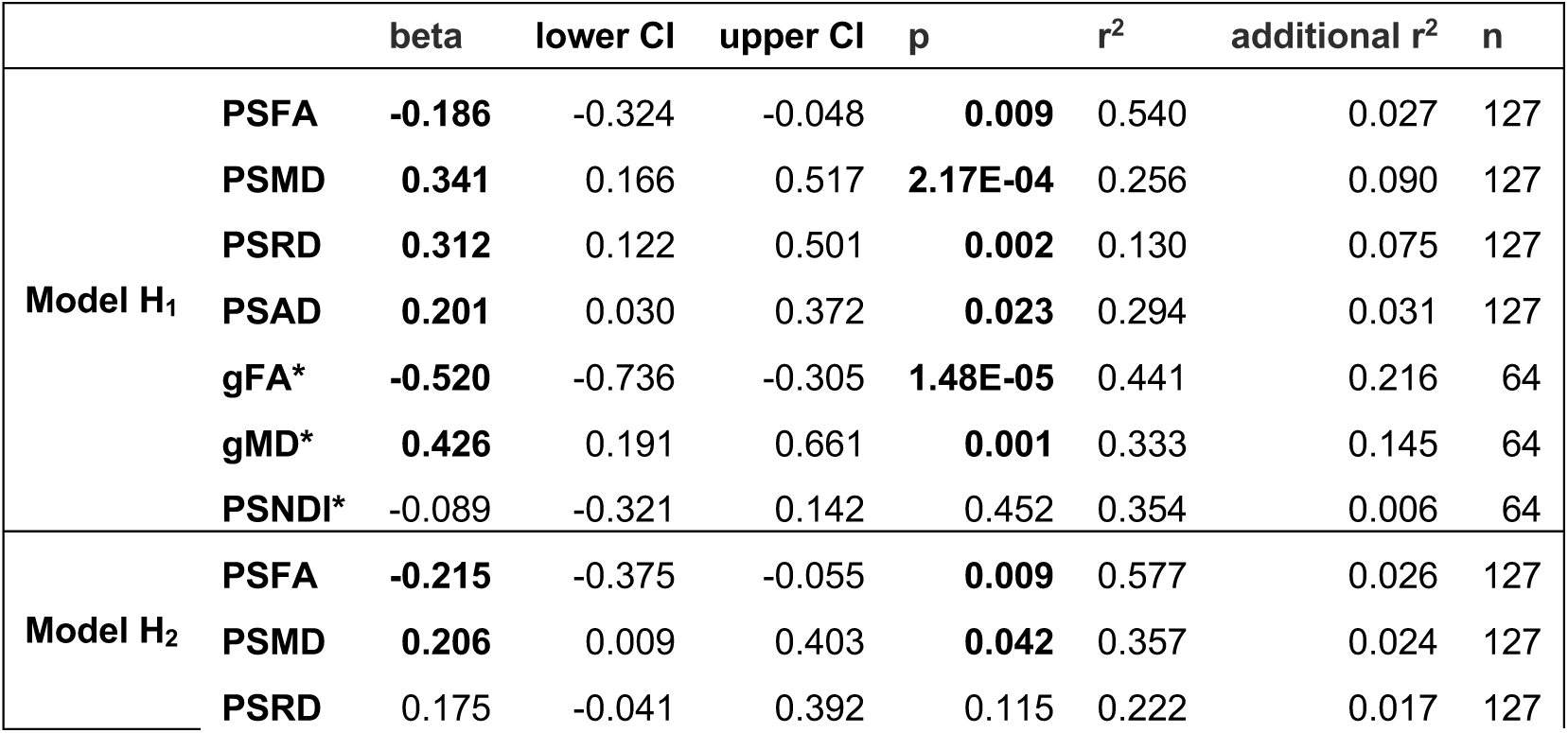
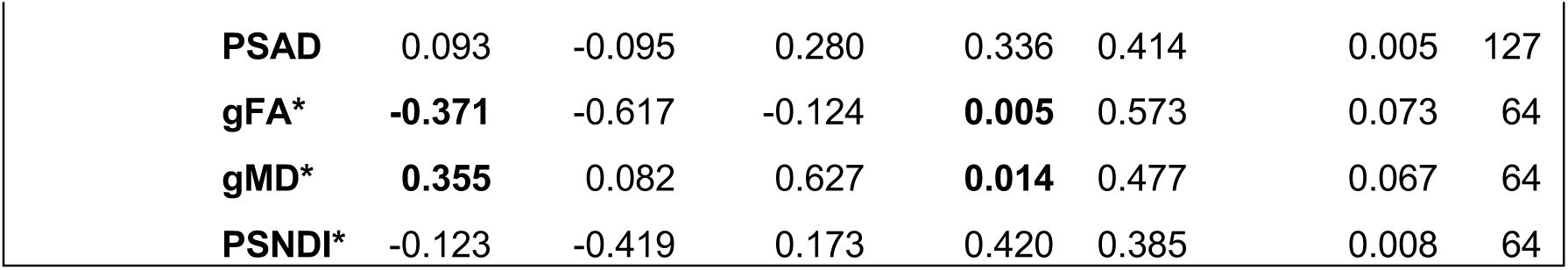
Associations between DNAm CRP and global white matter microstructure and the impact of inflammatory risk factors on associations in preterm infants; standardized regression coefficients for DNAm CRP associations between global white matter microstructure metrics for preterm infants. Betas (standardized coefficients) and 95% confidence intervals are reported. Bold text indicate statistically significant association (FDR-corrected p < 0.05). Model H_1_ controls for infant sex, gestational age at birth, gestational age at scan birthweight Z score and scanner variable; model H_2_ additionally controls for inflammatory risk factors and associated morbidities (maternal smoking in pregnancy, preeclampsia, HCA, sepsis, BPD, NEC and ROP) *scanner variable is controlled for when examining PS metrics but not gFA, gMD and PSNDI (single-scanner sample).

|  |  | beta | lower CI | upper CI | p | r<sup>2</sup> | additional r<sup>2</sup> | n |
| --- | --- | --- | --- | --- | --- | --- | --- | --- |
| Model H<sub>1</sub> | PSFA | **-0.186** | -0.324 | -0.048 | **0.009** | 0.540 | 0.027 | 127 |
|  | PSMD | **0.341** | 0.166 | 0.517 | **2.17E-04** | 0.256 | 0.090 | 127 |
|  | PSRD | **0.312** | 0.122 | 0.501 | **0.002** | 0.130 | 0.075 | 127 |
|  | PSAD | **0.201** | 0.030 | 0.372 | **0.023** | 0.294 | 0.031 | 127 |
|  | gFA* | **-0.520** | -0.736 | -0.305 | **1.48E-05** | 0.441 | 0.216 | 64 |
|  | gMD* | **0.426** | 0.191 | 0.661 | **0.001** | 0.333 | 0.145 | 64 |
|  | PSNDI* | -0.089 | -0.321 | 0.142 | 0.452 | 0.354 | 0.006 | 64 |
| Model H<sub>2</sub> | PSFA | **-0.215** | -0.375 | -0.055 | **0.009** | 0.577 | 0.026 | 127 |
|  | PSMD | **0.206** | 0.009 | 0.403 | **0.042** | 0.357 | 0.024 | 127 |
|  | PSRD | 0.175 | -0.041 | 0.392 | 0.115 | 0.222 | 0.017 | 127 |

**Table 2. Associations between DNAm CRP and global white matter microstructure and**
|  |  |  |  |  |  |  |  |
| --- | --- | --- | --- | --- | --- | --- | --- |
| **PSAD** | 0.093 | -0.095 | 0.280 | 0.336 | 0.414 | 0.005 | 127 |
| **gFA\*** | **-0.371** | -0.617 | -0.124 | **0.005** | 0.573 | 0.073 | 64 |
| **gMD\*** | **0.355** | 0.082 | 0.627 | **0.014** | 0.477 | 0.067 | 64 |
| **PSNDI\*** | -0.123 | -0.419 | 0.173 | 0.420 | 0.385 | 0.008 | 64 |

As with global brain volumetric measures, there were significant interactions between gestational age and DNAm CRP across measures of white matter integrity excepting PSRD: PSFA (interaction β = 0.225; main effect β = -0.212), PSMD (interaction β = -0.257; main effect β = 0.371) and PSAD (interaction β = -0.271; main effect β = 0.232), indicating infants at younger gestational ages were more likely to have poor white matter integrity with high DNAm CRP. When controlling for inflammatory risk factors, DNAm CRP associations between PSRD and PSAD were no longer significant.

Within a smaller subgroup of this sample, individual tract FA and MD as well as neurite density index data was available (Phase 2, refer to **supplementary figure 2** for study flow diagram). PCA-derived single-metric g-factors, gFA and gMD (scree plot and PCA variable contributions illustrated in **supplementary figure 6**) were almost exactly correlated with those previously reported in a larger sample of Theirworld Edinburgh Birth cohort infants (79). When examining the association between DNAm CRP and global white matter measures in this subsample, the most striking association was seen with differences in a general factor of fractional anisotropy, gFA (β = -0.52 [95% CI -0.304, -0.736], p = 1.48 × 10^−5^, incremental R^2^ = 22%) and mean diffusivity, gMD (β =0.423 [95% CI 0.661, 0.191], p = 7.79 × 10^−4^, incremental R^2^ = 14%). These effect sizes were attenuated by controlling for additional inflammatory risk factors (model H_2_) but remained significant (β range |0.35| to |0.37|, p < 0.05). No significant associations were found between DNAm CRP with PSNDI.

### 3.5 Individual white matter tract associations with DNAm CRP

We next examined associations between DNAm CRP and individual tract-averaged FA and MD. In all models, term infants displayed no significant tract associations with DNAm CRP (**supplementary figure 7)**. In preterm infants, altered FA was present in both hemispheric tracts of the AF, CST, IFOF, ILF, UNC and CCG (**Figure 4** shows tract-averaged fractional anisotropy for each of the 16 tracts for the term and preterm neonates). Some tract associations were specific to hemisphere such as decreased FA in the left (but not right) ATR. Equally, altered FA and MD was present in only the genu (and not splenium) of the corpus callosum. A breakdown of all dMRI results is reported in **supplementary table 11**. However, after adjusting for additional inflammatory risk factors, (in order of effect size) only FA in the right corticospinal tract (15.4%), right AF (10%), and right CCG (8.7%) remained significant. For tract MD, bilateral increases in MD were observed in the AF, CST, IFOF, ATR and CCG. Hemispheric specific associations were found for the left ILF, left ATR and genu of the corpus callosum. Of these associations, AF and ILF were no longer significant when accounting for additional inflammatory exposures.

**Figure 4.**
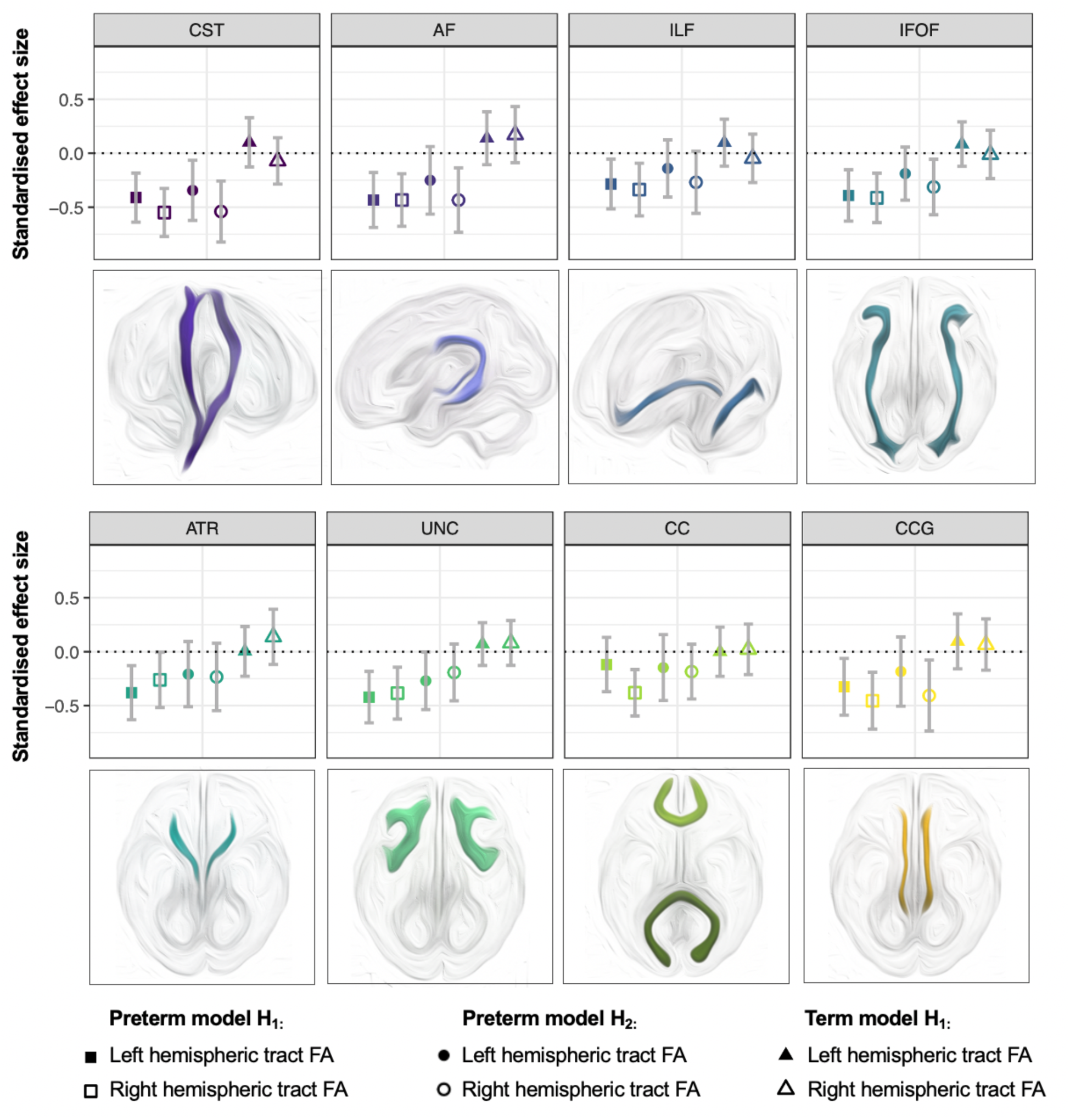
DTI-tract associations with DNAm CRP. Standardized regression coefficients for DNAm CRP associations between tract fractional anisotropy (FA) for preterm infants (squares), preterm infants in models controlling for additionally inflammatory risk factors (circles) and term infants (triangles). Filled shapes are left tracts and open shapes are right hemispheric tracts, except in the case of the CC where filled shapes are the splenium and open shapes are the genu of the corpus callosum. Points show standardized coefficients and 95% confidence intervals. All models are controlled for sex, gestational age at birth, gestational age at scan and birthweight Z score. Model H_2_ (circles) additionally controls for inflammatory risk factors (maternal smoking during pregnancy, preeclampsia, HCA, neonatal sepsis, BPD, NEC, ROP). For MD associations see **supplementary figure 7**.

## 4. Discussion

In this study, we integrated data from placenta, saliva and brain MRI in a large cohort of 258 infants to characterise the association of inflammation with brain structure. We demonstrate that a composite buccal-cell DNA methylation measure of inflammation trained in adult peripheral blood samples associates with comorbidities of preterm birth that are characterised by a pro-inflammatory state and widespread differences in brain structure among preterm infants. The epigenetic signature was particularly associated with white matter dysmaturation at term-equivalent age both globally and at the level of individual white matter tract microstructure, associations that largely remained significant when accounting for inflammatory exposures. Using this epigenetic signature which has previously tracked with inflammation and brain structural alterations in older age human cohorts, these data motivate further research into the potential of immune-DNAm markers for translational medicine in the neonatal period as diagnostic tools for identifying those at risk for inflammatory-related morbidities and neurodevelopmental impairment.

### 4.1 Inflammatory-related DNAm biomarkers

There has been increasing interest in using methylation data to advance our understanding of the causes and consequences of preterm birth as outlined in several reviews (10,88–91). Only recently has attention turned to exposures in the perinatal period, the role of epigenetics *in utero* for neurodevelopment, and the potential of peripherally sampled DNAm to capture the impact of environmental exposure in relation to brain and cognitive outcomes (92). Among these developments are the use of poly-epigenetic scores of exposure, which integrate information from multiple CpG sites to provide a record of exposure or to capture a complex trait (93). This approach has been used to examine maternal smoking (94–97) glucocorticoid, and prenatal folate exposure during pregnancy (55,98), alongside environmental exposures such as pollution (98). Examining DNAm proxies of inflammation is uncommon, but given the health associations with inflammatory-related DNAm in adult cohorts (47,48,99), and the shared nature of epigenetic changes between mother and infants (64,100), we hypothesised that variance in the neonatal methylome could reflect a convergence of amassed inflammatory burden from different perinatal origins.

### 4.2 DNAm CRP associates with gestational age and multiple inflammatory exposures

Our findings suggest that epigenetics offers a solution to the traditional limitations of assessing inflammatory burden in infancy. Preterm infants displayed higher DNAm CRP than term infants, and associations between DNAm CRP and postnatal health and brain outcomes were restricted to the preterm infants. This novel finding that gestational age at birth correlates strongly with epigenetic inflammation (r = -0.62, p <0.001) aligns with previous observations of elevated inflammatory protein concentrations with lower gestational ages and prematurity (8,15), lending further weight to the validity of this measure for carrying clinical significance. Equally, finding that inflammation-related alterations in brain structure were reserved to preterm infants (and no trends were seen in term infants) is likely because of the rapid developmental changes during the second and third trimester of pregnancy for both the developing innate immune system and brain – in particular, the disruptive impact of inflammation on neurogenesis, neuronal migration, synaptogenesis and myelination (3). As these processes are highly dynamic during these periods and early postnatal life, preterm infants are both more susceptible to sustained inflammation and neurodevelopmental disruption – the mechanisms of which we outline below.

Our finding that multiple inflammatory hits contributed to higher DNAm CRP strengthens the hypothesis that DNAm may index the allostatic load of inflammation during neonatal intensive care. In both preclinical studies and cohort groups, preterm infants with multiple inflammatory episodes or morbidities display an increased risk for brain structural abnormalities compared to infants who had only one inflammatory episode or condition recorded (101–103). The multi-hit hypothesis of sustained inflammation (14,26,101) suggests that postnatal health complications related to preterm birth can perpetuate a chronic inflammatory state, with timing of insults a key factor for why preterm infants are more susceptible than term infants to sustained inflammation (104). This could affect DNA methylation in the immune system, with studies demonstrating that lower birthweight infants go on to display higher concentrations of CRP in adolescence and adulthood (105,106), and that adversity-related changes in immune cell DNAm are related to raised plasma inflammatory mediators (107). Epigenetic modifications are an essential mechanism by which inflammatory risk factors could lead to long-term disruptions in both immune and brain development (54,108), and this work highlights the utility of profiling such changes alongside other clinical and biological data.

### 4.3 White and deep grey matter dysmaturation in preterms with elevated inflammation

The most striking finding from this study is the association of DNAm CRP with widespread variances in brain structure in preterm but not term infants (Figures 2-4). We observe larger effect sizes for associations of DNAm CRP with global white matter microstructure (gFA and gMD) than white matter volume in a sub-population of these infants. This may be because diffuse white matter injury antedates overall reductions in white matter volume, with DNAm CRP-DTI associations capturing a more subtle dysmaturation of programmed development (109). Both global white matter volume, microstructure and regional white matter integrity were lower in preterms with elevated DNAm CRP, with infants at younger gestational ages more prone to elevated inflammation and related poor white matter integrity. These findings echo the results of prior studies (25,33,110,111), and are overall consistent with the theory that alterations in white matter microstructure are largely a consequence of dysregulation of white matter development driven by inflammation (111).

In addition to white matter, volume reductions were observed in the hippocampi and amygdale and deep grey matter with increased DNAm CRP, though the former did not remain significant after accounting for additional inflammatory risk factors (**supplementary table 10**). The association of elevated DNAm CRP with lower deep grey matter volume is consistent with previous research that finds that preterms infants exhibit deep grey matter loss relative to term infants (112–114). Given the relationship we outline here between preterm birth and inflammatory load, inflammation may be a key driver of these differences, both via its direct effects on brain structure and its contribution to related damage such as sensitisation to hypoxia ischaemia, excitotoxic insults and other early-life stressors (2,115). These widespread alterations in brain structure are particularly interesting given the evidence base for inflammation relating to cognitive impairment, as studies have shown that both hippocampal volume and thalamic volume loss accompanying white matter microstructural alterations are linked to neurodevelopmental outcomes in early childhood (116–118). Inflammation-related grey matter loss is considered a consequence of dysregulated neuronal development, with inflammatory mediators disrupting processes such as dendritic arborization and cortico-thalamic connectivity (110). As consolidation of thalamocortical connections happens in the third trimester of pregnancy, deep grey matter structures may be vulnerable to inflammatory stimuli (119).

We also observed regional variance in how DNAm CRP associates with white matter tracts, a finding consistent with previous studies that indicate that certain white matter tracts are more vulnerable to inflammatory-adjacent events such as hypoxia ischemia (120,121) traumatic brain injury (9), intraventricular haemorrhage (122) and cerebral palsy (5). Different white matter tracts develop at different rates *in utero* and display distinct transient growth periods of increased axonal development. These windows of plasticity have been outlined as particularly vulnerable to perturbation (123,124), with inflammation disrupting the developmental lineage of oligodendrocytes, resulting in hypomyelinated axons (1,110,125,126). Developmental growth periods of certain white matter tracts may therefore underscore regional vulnerability to elevated inflammation, with younger tracts likely to have higher proportions of pre-myelinating oligodendrocytes vulnerable to inflammatory mediators. However, we caution that we lack the statistical power to reliably detect differences between the magnitude of associations in regional white matter structure, and instead interpret these findings as evidence of the pervasive and widespread impact of inflammation on the development of white matter. Correspondingly, though there were differences between the association significance for the left and right hemispheres for several of the delineated tracts, these unilateral findings are in keeping with previous studies of similar sample size (4,127); as the magnitudes were similar (with overlapping confidence intervals), this did not indicate a strong basis for laterality of effects.

### 4.4 Strengths and limitations

To our knowledge, this is the first time an epigenetic measure of inflammation has been examined in a preterm cohort in relation to brain health outcomes. The effect sizes reported in this study are consistent with that of previous epidemiologic studies of DNAm and early life outcomes (128). The sample size (n = 258 for inflammatory exposure, and n = 121-214 for neuroimaging associations), is akin to that of previous work examining inflammation and brain structure in preterm infant populations (6,11,36,37,129–131), and in many cases more substantial, with the vast majority of prior work conducted in sample sizes of less than 100 infants (4,19,21,32–35,127,132). There is a distinct scarcity of detailed methylation alongside multi-modal neuroimaging data (133,134), particularly in neonatal cohorts (61,62,135), making this a valuable contribution to the DNAm-neuroimaging field.

Although the weights for the predictor were trained in adult blood samples, we observed similar associations between DNAm CRP and brain structural outcomes to those in previous studies of adults (47,48). Given we have now applied this method to buccal-cell DNAm, it is encouraging to see similar associations between DNAm CRP and brain structural outcomes, especially given that DNAm is highly tissue specific (136), and previous research has reported on cross-tissue differences in magnitude and direction of effects for other traits (137). This cross-tissue approach (where weights were originally created from blood-based DNAm, and later applied to saliva-based composite signatures) has also been adopted in other studies (98,138). While future studies would ideally measure CRP directly from serum or blood spots in infants to enable direct cross-tissue comparisons, saliva has the advantage of being one of the most accessible tissue samples for infant populations, and may be more suitable than other peripheral samples (such as blood) when examining brain and cognitive outcomes owing to the brain and buccal cell shared ectodermal origins (139–142). In the absence of direct comparison with inflammatory mediators from blood samples, the strong correlates with clinical inflammatory conditions (both fetal and postnatal) is affirming, and we have taken steps to account for possible sources of confounding (**supplementary figure 1**).

Neuroimaging studies are notoriously heterogenous in their design given the array of different MRI acquisition techniques, processing pipelines and chosen outcome measures. The choice of neuroimaging features is even more relevant in the context of preterm birth to adequately address the motivating research questions (38). Here, our choice of neuroimaging features was guided by established characterizations of EoP in preterm infants, namely water content and dendritic/axonal complexity and dysmaturation within the white matter, and grey matter volume (77,124). While we consider this comprehensive characterisation of brain structure from NODDI and dMRI data a significant strength of this study, we acknowledge that microstructure measures such as FA and MD in older cohorts are commonly considered surrogates of white matter integrity or myelination, the white-matter pathways in this study are still developing at the time of gestational age at scan (range 37.70 - 45.14 weeks), and as such may not reflect permanent differences. Longitudinal follow up is therefore encouraged for future studies designed to examine the implications of sustained inflammation in preterms for neurodevelopmental outcomes and lifecourse brain health.

We do not attempt to discuss the causality of the relationship between DNAm CRP with brain structure, though the causality of such associations is a persistent topic of debate in epigenetic epidemiological research and has been discussed in depth in reviews (143). Future work examining transcriptomic changes on the same peripheral samples from which DNAm data is collected, as well as statistical approaches like two-step mendelian randomization, are important developments to unpick causality of these relationships. Studies investigating whether these differences in DNAm remain, amplify, or attenuate with age are advised, as well as how sensitive these signatures are in the context of intervention (anti-inflammatory medications and treatments, as well as lifestyle interventions such as the cessation of smoking in pregnancy). There is also precedent to examine whether composite methylation proxies of inflammation differ across psychopathology (144) or specific neurological cases such as Cerebral Palsy or neurodevelopmental disorders such as Fragile X syndrome, autism and ADHD, given examples of other poly-epigenetic signatures of psychiatric disorders (145). Future work that focuses on such DNAm dynamics in relation to these outcomes in ongoing longitudinal studies of infants born preterm is therefore of interest, as well as replication in different population samples.

Finally, DNAm was sampled in neonates postnatally. While this is rational when examining variances in DNAm and brain structural differences in infants, future studies that examine both maternal and infant DNAm could examine the degree to which exposures are shared or specific to parent and offspring. Equally, multiple DNAm sampling during pregnancy could elucidate key critical periods of susceptibility to inflammation by parsing out exposures specific to trimester or months of pregnancy, affording new insights into the spatiotemporal patterning of brain development in relation to dynamic immune changes in the perinatal period.

## 5. Conclusion

Inflammatory-related DNAm is associated with risk of postnatal health outcomes and brain dysmaturation. Our results indicate that multiple inflammatory-related hits from different origins (pertaining to maternal, fetal, and postnatal exposures) may be captured by changes to the DNA methylation profiles of infants and may help to explain variances in brain structure in preterm populations, circumventing limitations of traditional measures of inflammation. As early birth is associated with sudden change in immune-related risks, which coincide with the developing immune system and windows of neurodevelopmental plasticity, it is theorised that preterm infants are at greater risk of inflammation-related disruption of white matter. Our work here provides new layers to this theory, with epigenetic inflammation associating with diffuse and global brain – and particularly white matter – alterations in preterm but not term infants, indicating that sustained inflammation may be a key driver of neurodevelopmental disruption. In summary, the association of an epigenetic signature with inflammatory outcomes, and inflammation-relevant neural phenotypes, supports the use of methylation data in integrative, multimodal approaches toward disease stratification in the perinatal period.

## Data Availability

All data produced in the present study are available upon request to the authors

https://www.tebc.ed.ac.uk/2019/12/data-access-and-collaboration

## 7. Acknowledgements

Some of the participants were scanned in the University of Edinburgh Imaging Research MRI Facility at the Royal Infirmary of Edinburgh which was established with funding from The Wellcome Trust, Dunhill Medical Trust, Edinburgh and Lothians Research Foundation, Theirworld, The Muir Maxwell Trust and other sources. We thank Thorsten Feiweier at Siemens Healthcare for collaborating with dMRI acquisitions (Works-in-Progress Package for Advanced EPI Diffusion Imaging). The authors are grateful to the families who consented to take part in the study and to all the University’s imaging research staff for providing the infant scanning.

