## Supplementary_document for "Immuno-epigenetic signature derived in saliva associates with the encephalopathy of prematurity and perinatal inflammatory disorders"

---

**This document includes:**

- Supplementary Methods
- Supplementary Results
- Supplementary Figures 1-7
- Supplementary Tables 1-11

### Supplementary methods:

#### DNA methylation pre-processing

Raw intensity (.idat) files were read into R environment (version 3.4.4) using minfi. wateRmelon and minfi used for preprocessing, quality control and normalisation (1). The pfilter function in wateRmelon was used to exclude: samples with 1 % of sites with a detection p-value greater than 0.05; sites with beadcount 0.05. Cross hybridising probes and probes targeting single nucleotide polymorphisms with overall minor allele frequency ≥0.05 were also removed. Control probes were also removed. Samples were removed if there was a mismatch between predicted sex (minfi) and recorded sex (n = 3). Probes located on sex chromosomes were removed prior to analysis. Data from one of each twin pair was removed randomly (n=20). Data was danet normalised which includes background correction and dye bias correction (1). Saliva contains different cells types, including buccal epithelial cells and leukocytes. Epithelial cell proportions were estimated with epigenetic dissection of intra-sample heterogeneity with the reduced partial correlation method implemented in the R package EpiDISH (2). Prior to implementation of statistical models, β-values for CpG sites were adjusted (regressed as dependent variables) to remove batch effects using ComBat (3), where each BeadChip was considered to be one batch, and effects of estimated epithelial cell proportions.

#### dMRI pre-processing

Phase 1 dMRI acquisition were denoised using a Marchenko-Pastur-PCA-based algorithm (4); eddy current distortion and head movement were corrected using outlier replacement (5,6); bias field inhomogeneity correction was performed by calculating the bias field of the mean b0 volume and applying the correction to all the volumes (7). The processing for phase 2 was very similar as the one for phase 1. The main differences are that because the phase 2 consists in two different acquisitions, the data was concatenated before starting the pre-processing, then due to the availability of reverse encoded data, we corrected for EP distortion and within volume movement (5,8).

#### Tract segmentation and extraction of tract-averaged dMRI metrics

The details for white matter tract segmentation are comprehensively outlined in a previous study by this group (9,10). Briefly, the ENA50 neonatal template space (brain atlas optimised for infants) was used to perform whole brain tractography (11) and the SingleTensorFT tool within DTI-TK (12) was used to parse out white matter tractography from the ENA50 atlas tensor volume. From here, segmentation of white matter tracts was performed within the ENA50 atlas and tracts were delineated by drawing regions of interest (ROIs) manually on the FA image, using the protocols outlined in (9,10).

### Supplementary results:

#### Bivariate analysis of potential covariates with DNAm CRP

Prior to assessing the relationships between DNAm CRP with brain structural outcomes or postnatal inflammatory morbidities, we explored the interrelationships between DNAm CRP and the other variables in our study (**supplementary figure 3**). Correlations were assessed with Pearson correlations between continuous variables (e.g. association between DNAm CRP scores and gestational age) and point-biserial correlations between continuous variables and binary variables (e.g. DNAm CRP scores with incidence of sepsis). Preterm status and DNAm CRP score were moderately correlated ($r$ (256) = .57, p > 0.001), and DNAm CRP scores were inversely associated with gestational age ($r$ (256) = -.62, p > 0.001). There were moderately strong correlations with risk of BPD (.5) moderate correlations with incidences of HCA and sepsis (.34-.37) and weak correlations with NEC, ROP and maternal smoking (.2-.28). There were no significant correlations between DNAm CRP and infant sex, birthweight Z score or preeclampsia but these were included in models due to biological significance (see DAG; **supplementary figure 1**). There were no significant correlations between DNAm CRP and maternal age or gestational diabetes, so these covariates were omitted from regression models.

### Supplementary figures:

#### Supplementary Figure 1: Directed acyclic graph (DAG)

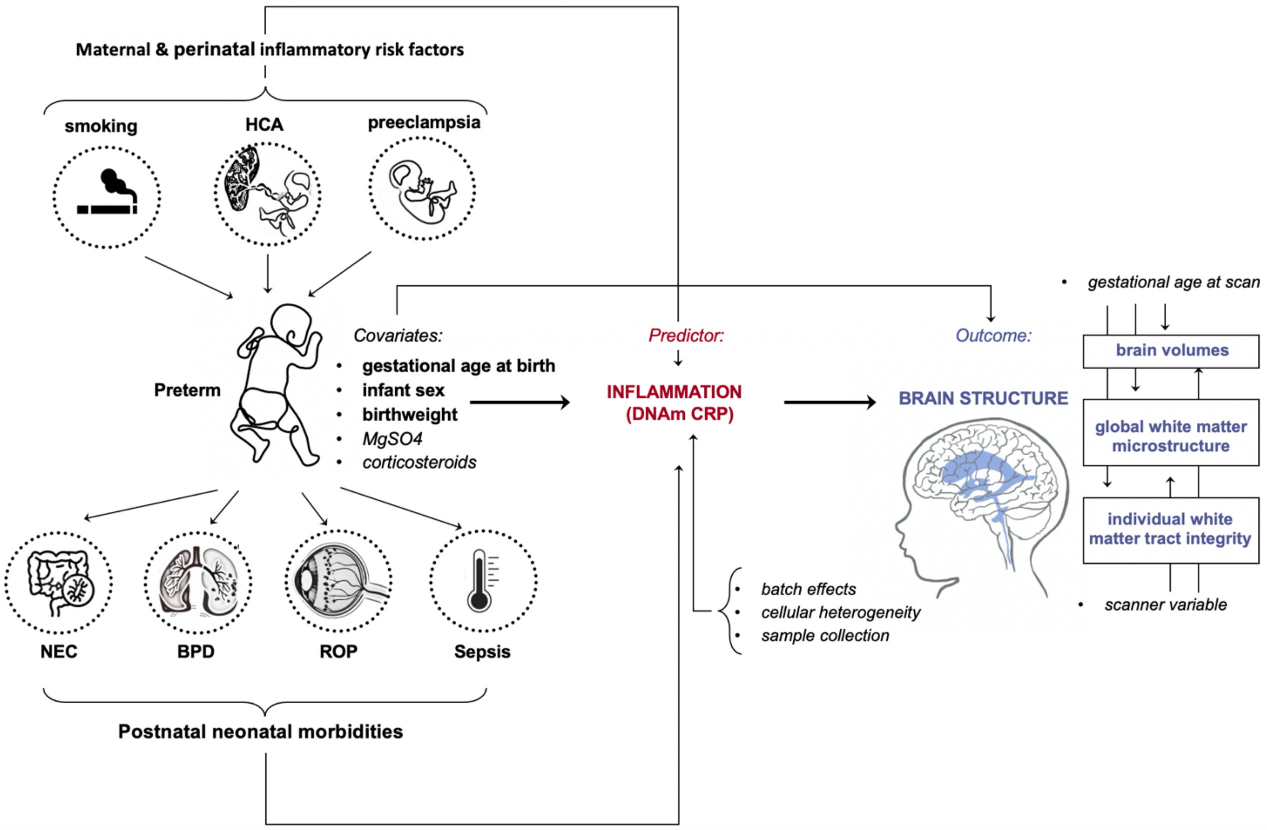


**Supplementary Figure 1:** Directed acyclic graph (DAG) demonstrating the expected interrelationships between preterm birth, DNAm CRP and brain structure. Covariates are in bold and black font and include maternal and perinatal inflammatory risk factors: smoking in pregnancy, preeclampsia and histologic chorioamnionitis (HCA); postnatal neonatal inflammatory morbidities, including bronchopulmonary dysplasia (BPD), necrotising enterocolitis (NEC), sepsis, and retinopathy of prematurity (ROP) as well as administration of MgS04 and corticosteroids in pregnancy. The predictor (DNAm CRP score, measured at same time as MRI scan), potential confounders (sex, gestational age at birth, birthweight, time of buccal swab collection), batch variables, cellular heterogeneity, and outcome variables (brain structural metrics). Variables that were adjusted for within models have are in bold; MgSO4 and corticosteroids was not adjusted for in all models, since it may result in over-adjustment, but was included for prediction of postnatal inflammatory conditions.

#### Supplementary Figure 2. Flow chart of study sample and data acquisition

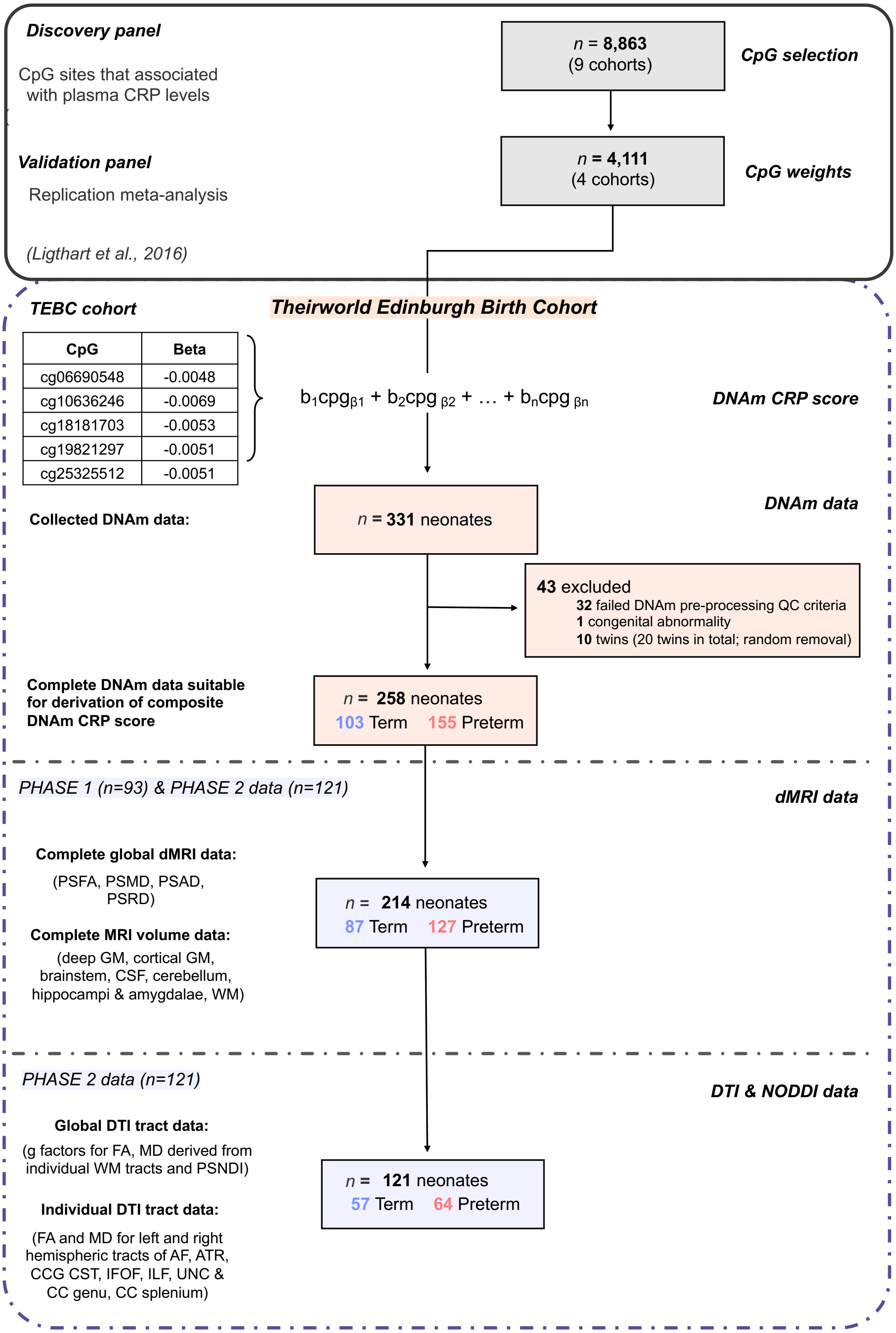


**Supplementary Figure 2. Flow chart of study sample and data acquisition**

Supplementary Figure 3. Correlations between study variables (n = 258)

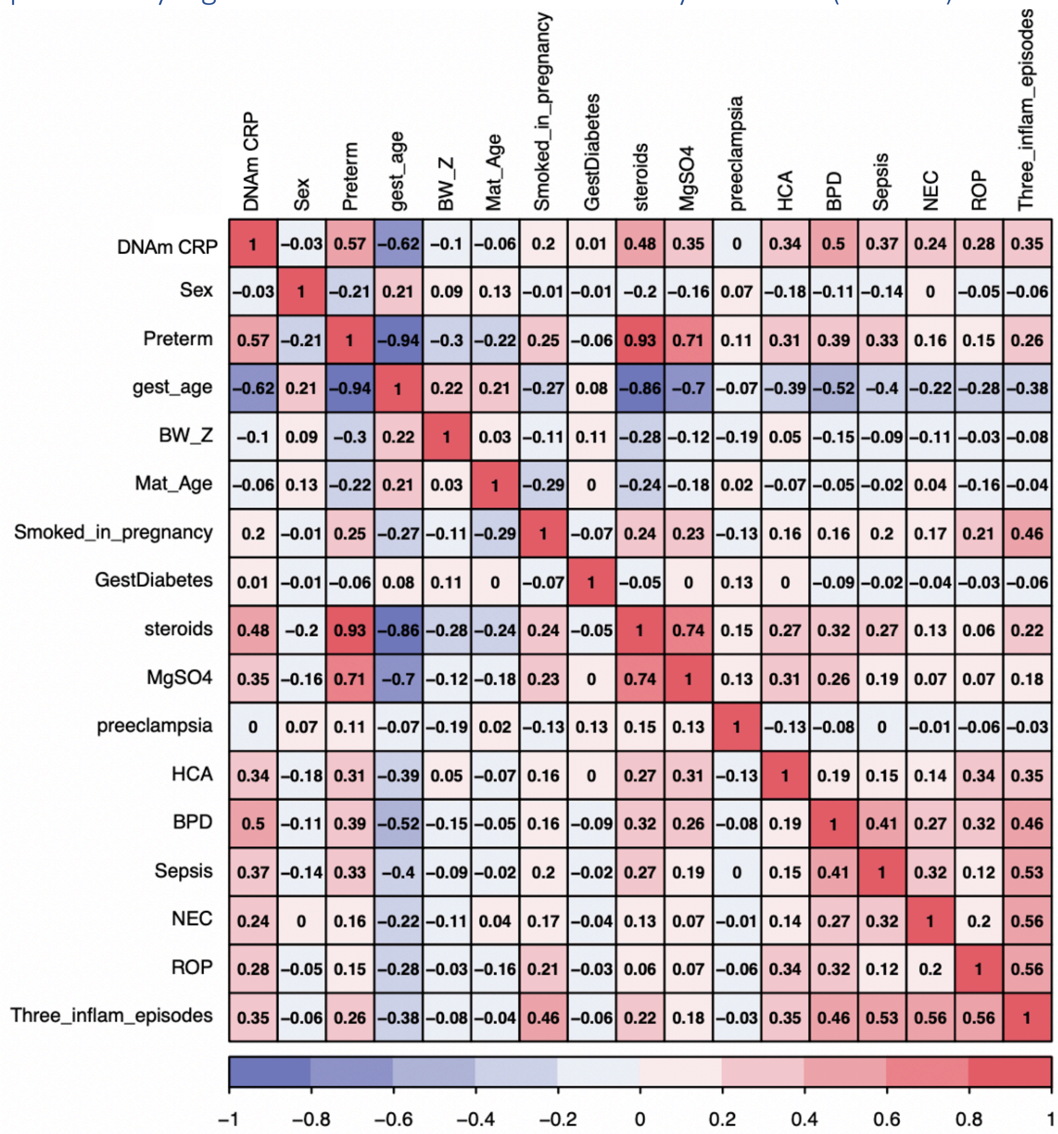


**Supplementary Figure 3. Correlations between study variables (n = 258)**

#### Supplementary Figure 4. model performance metrics for the association of DNAm CRP with white matter volume

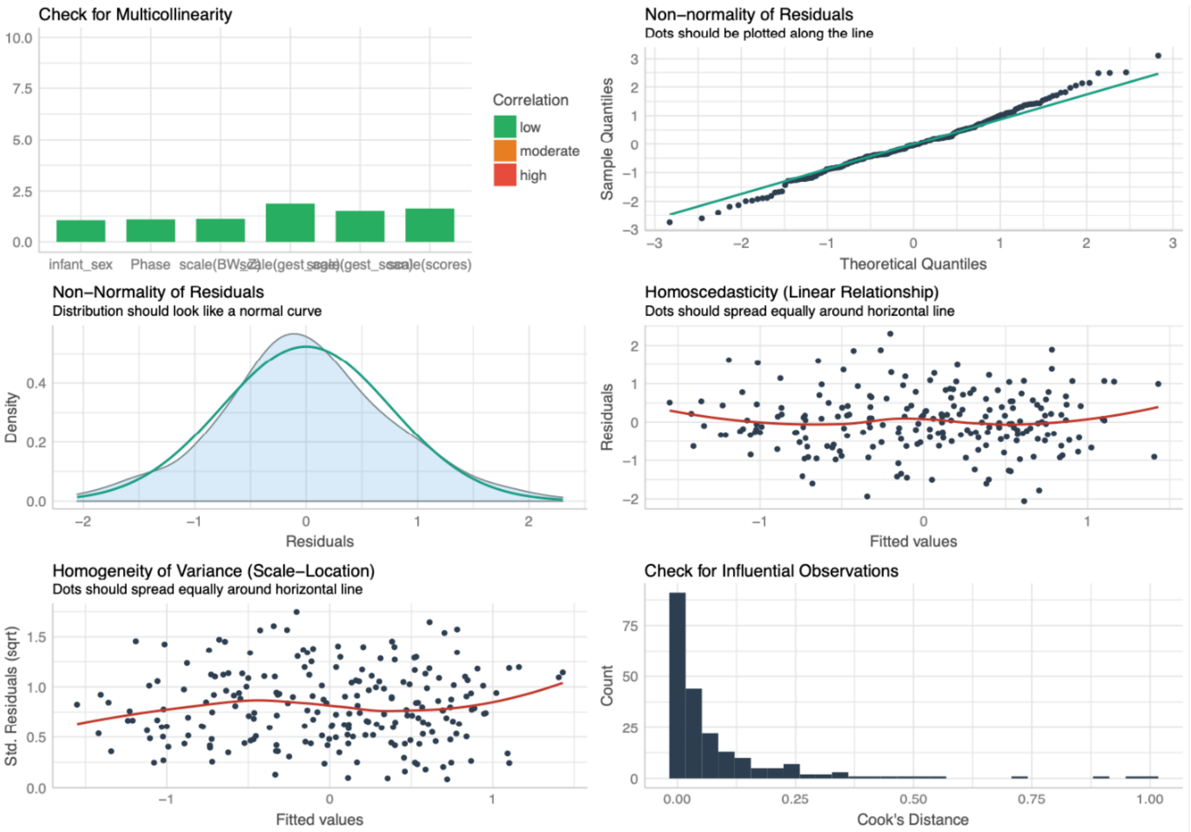


**Supplementary Figure 4.** Example model performance for *white matter volume ~ DNAm CRP + gestational age at birth + gestational age at scan + scanner variable + infant sex + birthweight (Z score)*

#### Supplementary Figure 5. Raw weighted FA and MD within sample

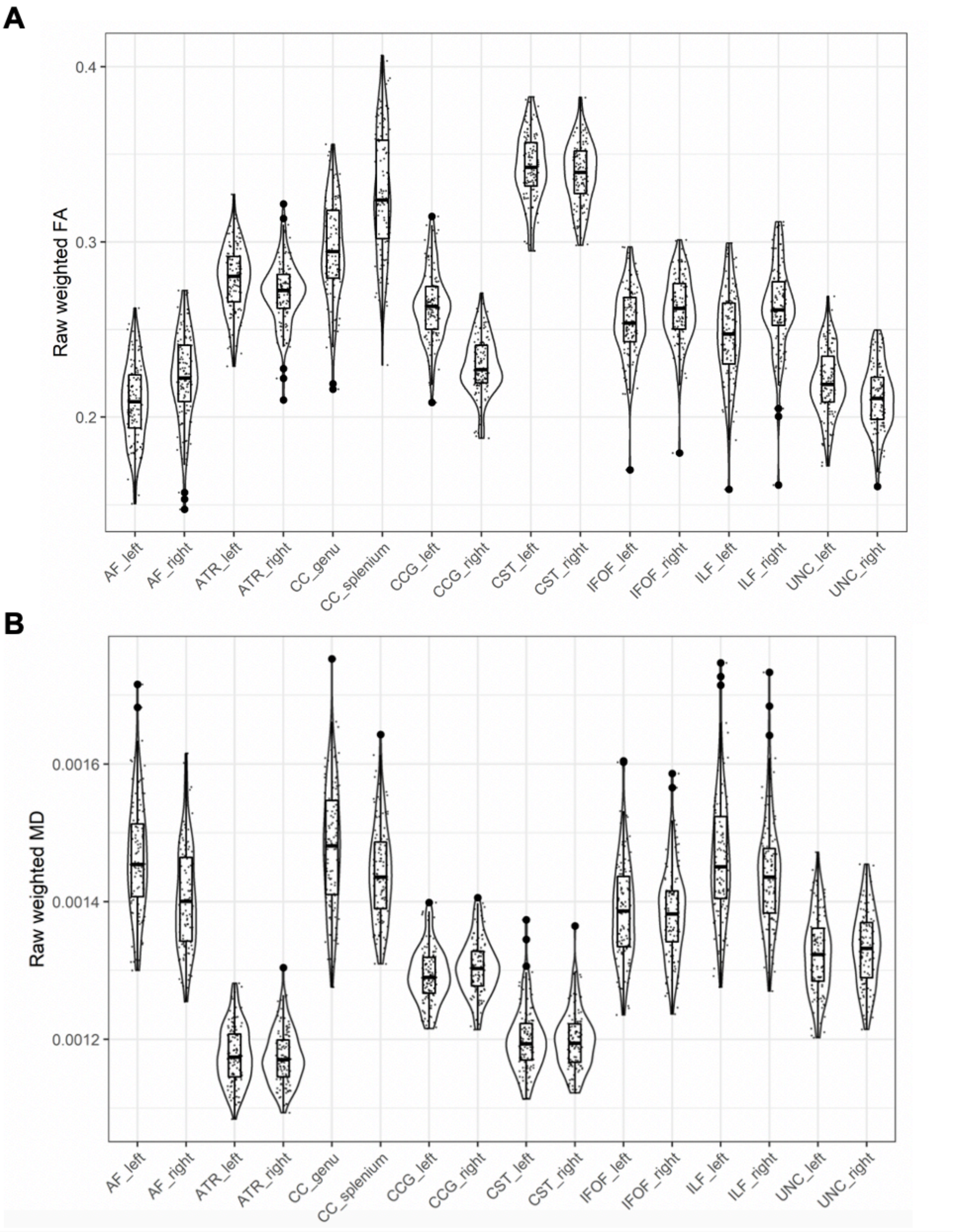


**Supplementary Figure 5. Raw weighted FA and MD values within subsample (n = 121)**

#### Supplementary Figure 6. Derivation of general factors of FA and MD

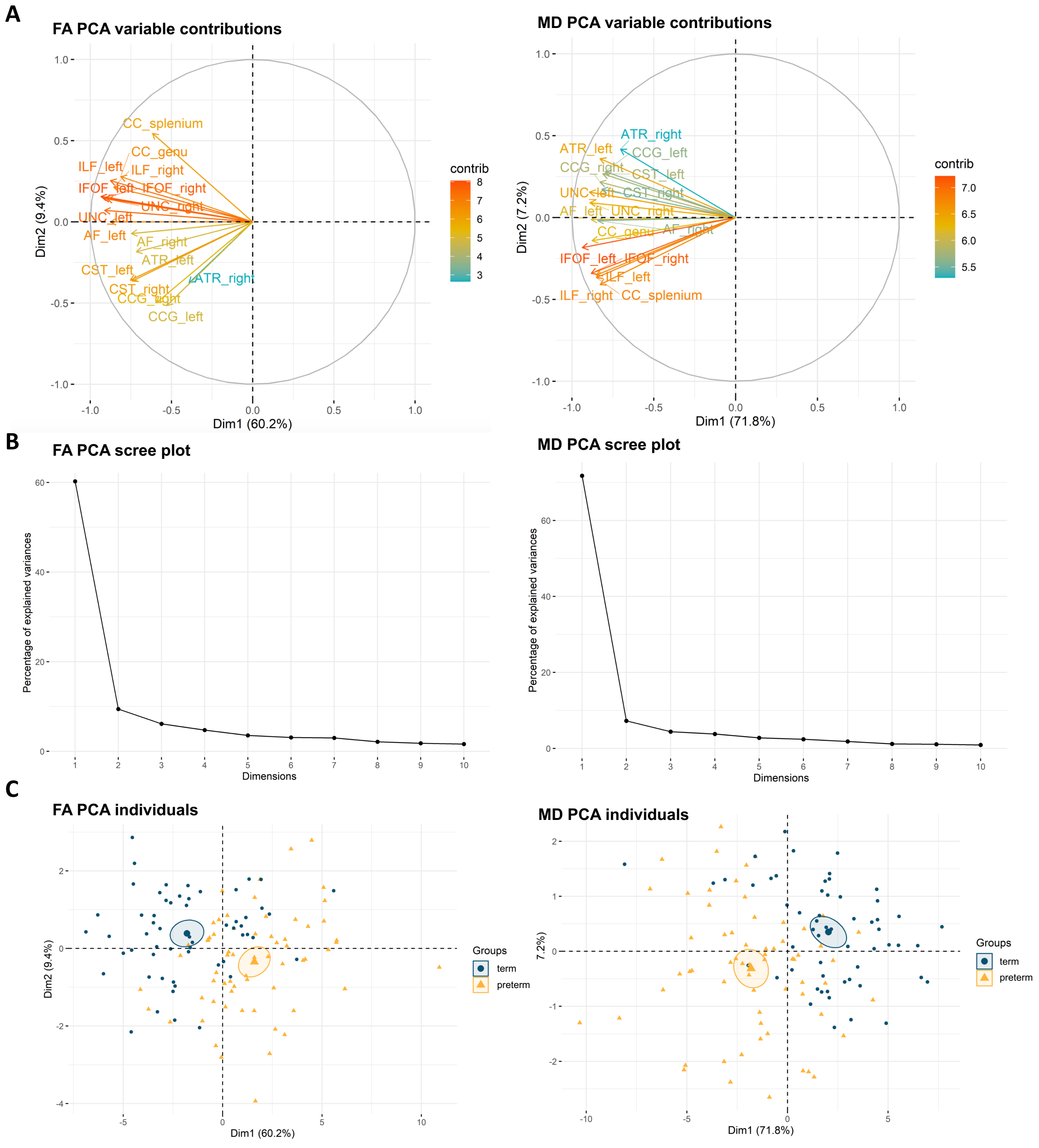


**Supplementary Figure 6. Derivation of general factors of FA and MD**. (A) PCA variable contribution plot; the colours represent the contribution of the dMRI metric to the components (B) Scree plot of the eigenvalues (C) visualisation of gFA and gMD between term (blue circles) and preterm (yellow triangles) on the multimodal principal component axes FA = fractional anisotropy, MD = mean diffusivity.

#### Supplementary Figure 7. DTI-tract associations with DNAm CRP

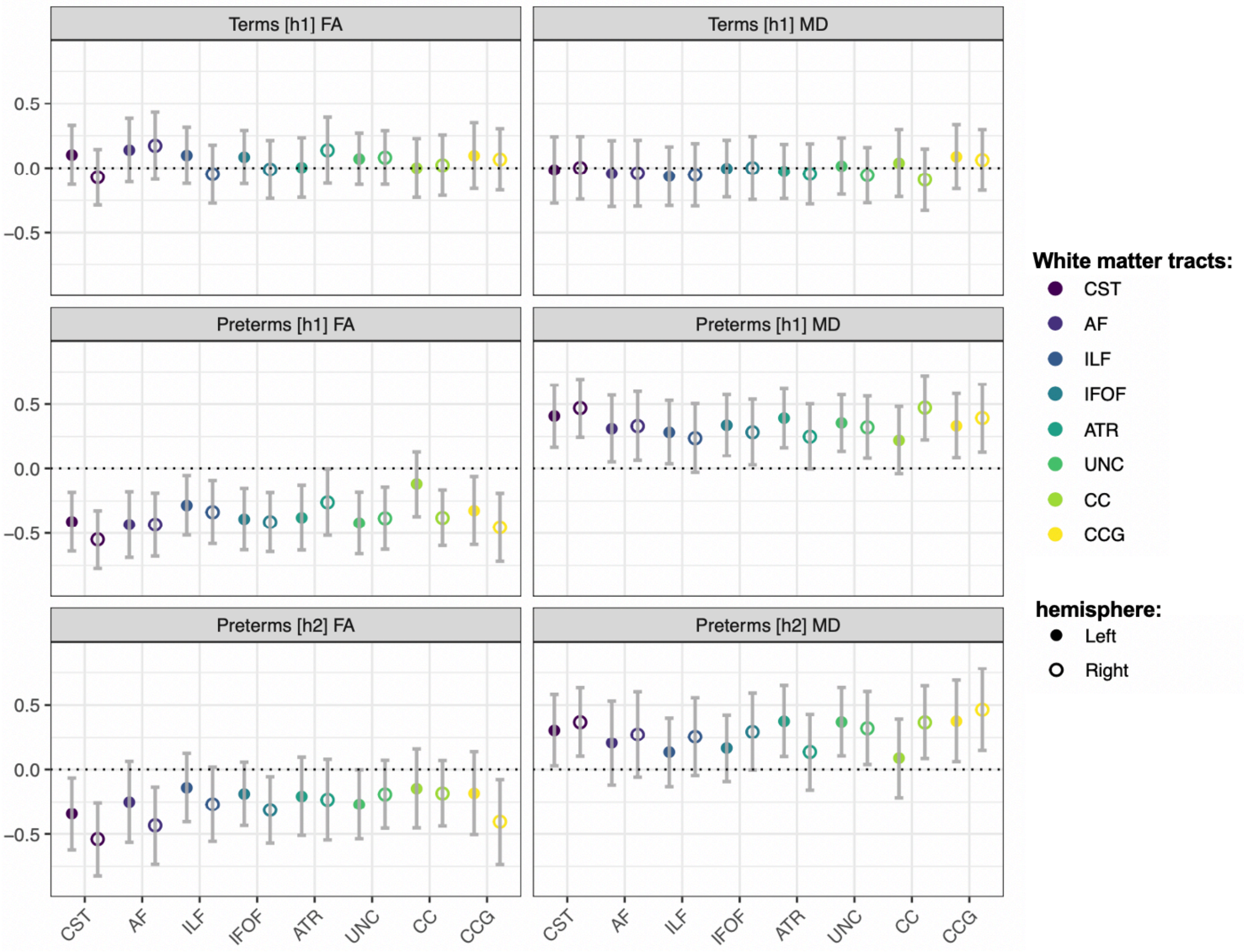


**Supplementary Figure 7. DTI-tract associations with DNAm CRP**. Standardized regression coefficients for DNAm CRP associations between tract fractional anisotropy (FA) [left] and mean diffusivity [right] for term (first row) preterm (second row) and preterm infants in models controlling for additionally inflammatory risk factors (third row). Filled circles are left hemispheric tracts and open shapes are right hemispheric tracts, except in the case of the CC where filled shapes are the splenium and open shapes are the genu of the corpus callosum. Points show standardized coefficients and 95% confidence intervals. All models are controlled for sex, gestational age at birth, gestational age at scan and birthweight Z score. Model $H_2$ [second row] additionally controls for inflammatory risk factors (which included incidence of smoking during pregnancy, preeclampsia, HCA, sepsis, BPD, NEC and ROP).

### Supplementary Tables

#### Supplementary table 1. CpG sites and relative weights

| *CpG* | *Gene* | **Beta (discovery sample)** |
|---|---|---|
| cg06690548 | ***SLC7A11*** | -0.0048 |
| cg10636246 | ***AIM 2 & IF116*** | -0.0069 |
| cg18181703 | ***SOCS3*** | -0.0053 |
| cg19821297 | ***DNASE2*** | -0.0051 |
| cg25325512 | ***FGD2*** | -0.0051 |
| cg27023587 | *HEATR6* | -0.005 |
| cg06126421 | *TUBB* | -0.0052 |

**Supplementary Table 1. CpG sites and relative weights** (from Lighthart et al. 2016) used to generate DNAm CRP score

#### Supplementary table 2. Cell-type expression of CpG sites used in composite methylation score

| | **macrophage** | **conventional dendritic cell** | **memory B cell** | **CD4-positive, alpha-beta T cell** | **CD8-positive, alpha-beta T cell** | **regulatory T cell** | **erythroblast** | **mature neutrophil** | **endothelial cell of umbilical vein (proliferating)** | **endothelial cell of umbilical vein (resting)** |
|---|---|---|---|---|---|---|---|---|---|---|
| *AIM2* | | 17 | 41 | | | 4 | | 3 | | |
| *DNASE2* | 59 | 1 | 8 | 5 | 3 | 11 | 15 | 2 | 11 | 14 |
| *FGD2* | 3 | 30 | 24 | | | | 2 | | | |
| *HEATR6* | 5 | 10 | 8 | 4 | 2 | 3 | 4 | | 5 | 5 |
| *IER3* | 16 | 80 | | 2 | 5 | 0.6 | 21 | 152 | 30 | 41 |
| *IFI16* | 22 | 20 | 40 | 21 | 18 | 38 | 9 | 16 | 44 | 51 |
| *SLC7A11* | 2 | 185 | | | | | 51 | 1 | 62 | 49 |
| *SOCS3* | 0.7 | 49 | 30 | 57 | 18 | 28 | 3 | 124 | 9 | 12 |
| *TUBB* | 128 | 145 | 34 | 40 | 81 | 49 | 366 | 33 | 311 | 316 |

**Supplementary Table 2. Peripheral expression of CpG sites within DNAm CRP score in tissue types**. Strand-specific RNA-Seq from different cell types from healthy individuals in the BLUEPRINT epigenome project. Light blue box: expression level is low (between 0.5 to 10 FPKM or 0.5 to 10 TPM); Medium blue box: expression level is medium (between 11 to 1000 FPKM or 11 to 1000 TPM); white: there is no data available. This study makes use of data generated by the Blueprint Consortium. A full list of the investigators who contributed to the generation of the data is available fromwww.blueprint-epigenome.eu.

#### Supplementary table 3. Association between DNAm CRP and inflammatory exposures

| outcome | model | OR | Lower CI | Upper CI | p value |
|---|---|---|---|---|---|
| | $H_1$ | **0.62** | 0.38 | 0.97 | 0.042 |
| preeclampsia | $H_2$ | **0.61** | 0.37 | 0.98 | 0.045 |
| | $H_3$ | 0.65 | 0.38 | 1.08 | 0.098 |
| | $H_1$ | 1.22 | 0.81 | 1.87 | 0.344 |
| smoked in pregnancy | $H_2$ | 1.20 | 0.79 | 1.84 | 0.398 |
| | $H_3$ | 1.32 | 0.84 | 2.11 | 0.237 |
| | $H_1$ | **1.87** | 1.29 | 2.78 | 1.29E-03 |
| HCA | $H_2$ | **1.95** | 1.32 | 2.97 | 1.21E-03 |
| | $H_3$ | **2.00** | 1.32 | 3.16 | 1.83E-03 |
| | $H_1$ | **3.29** | 2.11 | 5.44 | 7.07E-07 |
| BPD | $H_2$ | **3.77** | 2.36 | 6.44 | 1.97E-07 |
| | $H_3$ | **4.71** | 2.74 | 8.87 | 1.90E-07 |
| | $H_1$ | **2.10** | 1.38 | 3.34 | 9.16E-04 |
| Sepsis | $H_2$ | **2.33** | 1.50 | 3.79 | 3.20E-04 |
| | $H_3$ | **2.30** | 1.46 | 3.81 | 6.36E-04 |
| | $H_1$ | **3.10** | 1.39 | 8.58 | 0.013 |
| NEC | $H_2$ | **3.35** | 1.48 | 9.57 | 9.67E-03 |
| | $H_3$ | **3.86** | 1.58 | 12.03 | 8.05E-03 |
| | $H_1$ | 0.64 | 0.35 | 1.13 | 0.135 |
| Term control (0 inflammatory episodes) | $H_2$ | 0.68 | 0.37 | 1.20 | 0.196 |
| | $H_3$ | 0.67 | 0.37 | 1.20 | 0.199 |
| | $H_1$ | 0.45 | 0.29 | 0.66 | 8.76E-05 |
| Preterms (0 inflammatory episode) | $H_2$ | 0.42 | 0.27 | 0.63 | 4.90E-05 |
| | $H_3$ | 0.38 | 0.24 | 0.59 | 3.05E-05 |
| | $H_1$ | 0.91 | 0.65 | 1.26 | 0.560 |
| Preterms (1 inflammatory episode) | $H_2$ | 0.91 | 0.66 | 1.27 | 0.586 |
| | $H_3$ | 0.92 | 0.65 | 1.30 | 0.651 |
| | $H_1$ | 1.54 | 1.01 | 2.42 | 0.052 |
| Preterms (2 inflammatory episode) | $H_2$ | **1.60** | 1.04 | 2.53 | 0.038 |
| | $H_3$ | **1.73** | 1.08 | 2.86 | 0.026 |
| | $H_1$ | **2.78** | 1.63 | 5.20 | 5.01E-04 |
| Preterms (3 inflammatory episode) | $H_2$ | **2.83** | 1.65 | 5.33 | 4.58E-04 |
| | $H_3$ | **3.06** | 1.72 | 6.05 | 4.31E-04 |

**Supplementary Table 3. Association between DNAm CRP and inflammatory exposures.** Results presented as mean (standard deviation) unless specified. CI Confidence interval, OR Odds ratio, model $H_1$ = DNAm CRP only; model $H_2$ = adjusted for gestation at sample collection, birthweight Z score, infant sex; model $H_2$ = adjusted for gestation at sample collection, birthweight Z score, infant sex and corticosteroid and MgS04 administration during pregnancy

#### Supplementary table 4. Association between DNAm CRP and inflammatory exposure categories

| group1 | group2 | estimate | Lower CI | Upper CI | p.adj | p.adj.signif |
|---|---|---|---|---|---|---|
| [Term] 0 hits | [Term] 1 hit | 0.000236 | -3.50E-04 | 0.000822 | 8.56E-01 | ns |
| [Term] 0 hits | [Preterm] 0 hits | 0.000654 | 2.75E-04 | 0.001033 | 1.92E-05 | **** |
| [Term] 0 hits | [Preterm] 1 hit | 0.001058 | 7.17E-04 | 0.001399 | 7.59E-14 | **** |
| [Term] 0 hits | [Preterm] 2 hits | 0.001381 | 9.45E-04 | 0.001817 | 7.31E-14 | **** |
| [Term] 0 hits | [Preterm] 3 or more hits | 0.001689 | 1.22E-03 | 0.002158 | 6.95E-14 | **** |
| [Term] 1 hit | [Preterm] 0 hits | 0.000418 | -2.09E-04 | 0.001044 | 3.95E-01 | ns |
| [Term] 1 hit | [Preterm] 1 hit | 0.000822 | 2.17E-04 | 0.001426 | 1.67E-03 | ** |
| [Term] 1 hit | [Preterm] 2 hits | 0.001145 | 4.82E-04 | 0.001807 | 1.89E-05 | **** |
| [Term] 1 hit | [Preterm] 3 or more hits | 0.001453 | 7.69E-04 | 0.002138 | 6.06E-08 | **** |
| [Preterm] 0 hits | [Preterm] 1 hit | 0.000404 | -2.88E-06 | 0.000811 | 5.29E-02 | ns |
| [Preterm] 0 hits | [Preterm] 2 hits | 0.000727 | 2.38E-04 | 0.001216 | 3.99E-04 | *** |
| [Preterm] 0 hits | [Preterm] 3 or more hits | 0.001035 | 5.17E-04 | 0.001554 | 4.20E-07 | **** |
| [Preterm] 1 hit | [Preterm] 2 hits | 0.000323 | -1.37E-04 | 0.000783 | 3.37E-01 | ns |
| [Preterm] 1 hit | [Preterm] 3 or more hits | 0.000632 | 1.40E-04 | 0.001123 | 3.71E-03 | ** |
| [Preterm] 2 hits | [Preterm] 3 or more hits | 0.000309 | -2.53E-04 | 0.00087 | 6.14E-01 | ns |

**Supplementary Table 4. Association between DNAm CRP and inflammatory exposure risk categories.** Post-hoc testing (Tukey) of inflammatory exposure categories.

#### Supplementary Table 5. Associations between DNAm CRP and brain structure in full cohort n=214

| | Term & Preterm infants (n=214) | | | | | Preterm infants (n=127) | | | | | Term infants (n=87) | | | | |
|---|---|---|---|---|---|---|---|---|---|---|---|---|---|---|---|
| | **b** | **Upper CI** | **Lower CI** | **p** | **r2** | **b** | **Upper CI** | **Lower CI** | **p** | **r2** | **b** | **Upper CI** | **Lower CI** | **p** | **r2** |
| **Cortical_grey_matter** | -0.102 | -0.047 | -0.157 | 0.064 | 0.636 | -0.118 | -0.255 | 0.018 | 0.091 | 0.551 | -0.010 | 0.077 | -0.097 | 0.907 | 0.474 |
| **Deep_grey_matter** | **-0.143** | -0.088 | -0.198 | **0.010** | 0.632 | **-0.198** | -0.331 | -0.066 | **0.004** | 0.575 | 0.024 | 0.118 | -0.070 | 0.799 | 0.387 |
| **White_matter** | **-0.219** | -0.150 | -0.288 | **0.002** | 0.432 | **-0.218** | -0.389 | -0.048 | **0.013** | 0.298 | -0.082 | 0.016 | -0.181 | 0.405 | 0.326 |
| **Hippocampi_and_Amygdala** | -0.133 | -0.064 | -0.201 | 0.055 | 0.429 | **-0.185** | -0.343 | -0.027 | **0.024** | 0.395 | 0.128 | 0.230 | 0.026 | 0.211 | 0.280 |
| **Cerebellum** | -0.100 | -0.042 | -0.158 | 0.084 | 0.599 | -0.145 | -0.289 | -0.001 | 0.050 | 0.501 | 0.030 | 0.109 | -0.049 | 0.708 | 0.566 |
| **CSF** | -0.018 | 0.043 | -0.079 | 0.770 | 0.551 | 0.001 | -0.137 | 0.139 | 0.989 | 0.542 | -0.074 | 0.004 | -0.153 | 0.348 | 0.571 |
| **Ventricles** | 0.127 | 0.211 | 0.043 | 0.131 | 0.149 | 0.067 | -0.127 | 0.261 | 0.500 | 0.087 | -0.091 | 0.020 | -0.202 | 0.413 | 0.144 |
| **Brainstem** | 0.001 | 0.068 | -0.066 | 0.987 | 0.452 | -0.062 | -0.241 | 0.118 | 0.502 | 0.243 | 0.099 | 0.194 | 0.004 | 0.300 | 0.372 |
| **PSFA** | **-0.236** | -0.168 | -0.303 | **0.001** | 0.451 | **-0.186** | -0.324 | -0.048 | **0.009** | 0.540 | -0.071 | 0.016 | -0.158 | 0.419 | 0.469 |
| **PSMD** | **0.130** | 0.193 | 0.067 | **0.040** | 0.522 | **0.341** | 0.166 | 0.517 | **2.17E-04** | 0.256 | -0.030 | 0.079 | -0.139 | 0.782 | 0.171 |
| **PSAD** | 0.062 | 0.136 | -0.011 | 0.398 | 0.344 | **0.201** | 0.030 | 0.372 | **0.023** | 0.294 | -0.024 | -0.144 | -0.363 | 0.230 | 0.167 |
| **PSRD** | 0.094 | 0.161 | 0.027 | 0.161 | 0.458 | **0.312** | 0.122 | 0.501 | **0.002** | 0.130 | 0.006 | 0.113 | -0.102 | 0.959 | 0.192 |

**Supplementary Table 5. Associations between DNAm CRP and brain structure**. Standardized betas (β) and P-values are reported from regression models where DNAm CRP is regressed onto MRI measures for all infants (first column, n=214) , preterm infants only (second column, n=127), and term infants only (first column, n=87), covarying for gestational age, sex, birthweight Z score, gestational age at scan, scanner variable (volumetric data are also corrected for head size). Additional R2 refers to the amount of variance in MRI measures accounted for DNAm CRP, beyond covariates. Bold text denotes FDR q-value <0.05.

#### Supplementary Table 6. Interaction effects of gestational age on DNAm CRP in full cohort (n=214)

| | **DNAm CRP** | | | | **DNAm_CRP x gestational age** | | | | | |
|---|---|---|---|---|---|---|---|---|---|---|
| | **b** | **Upper CI** | **Lower CI** | **p** | **b** | **Upper CI** | **Lower CI** | **p** | **r2** | **add r2** |
| Cortical_grey_matter | -0.067 | -0.006 | -0.128 | 0.273 | 0.080 | 0.140 | 0.021 | 0.180 | 0.639 | 0.009 |
| Deep_grey_matter | -0.071 | -0.010 | -0.131 | 0.242 | **0.163** | 0.222 | 0.104 | **0.006** | 0.645 | 0.025 |
| White_matter | **-0.152** | -0.076 | -0.227 | **0.046** | **0.152** | 0.226 | 0.078 | **0.041** | 0.444 | 0.039 |
| Hippocampi_and_Amygdala | -0.082 | -0.005 | -0.158 | 0.285 | 0.115 | 0.190 | 0.041 | 0.123 | 0.435 | 0.017 |
| Cerebellum | -0.054 | 0.010 | -0.117 | 0.401 | 0.105 | 0.168 | 0.043 | 0.094 | 0.604 | 0.011 |
| CSF | -0.026 | 0.042 | -0.094 | 0.698 | -0.019 | 0.047 | -0.086 | 0.774 | 0.551 | 0.000 |
| Ventricles | 0.099 | 0.192 | 0.006 | 0.290 | -0.064 | 0.028 | -0.155 | 0.485 | 0.151 | 0.011 |
| Brainstem | 0.055 | 0.130 | -0.019 | 0.457 | 0.123 | 0.196 | 0.050 | 0.093 | 0.460 | 0.007 |
| PSFA | -0.111 | -0.038 | -0.183 | 0.127 | **0.283** | 0.354 | 0.212 | **8.88E-05** | 0.491 | 0.072 |
| PSMD | **0.131** | 0.201 | 0.061 | **0.064** | 0.002 | 0.070 | -0.067 | 0.981 | 0.522 | 0.010 |
| PSAD | -0.047 | 0.033 | -0.128 | 0.555 | **-0.248** | -0.170 | -0.327 | **0.002** | 0.374 | 0.033 |
| PSRD | 0.144 | 0.218 | 0.070 | 0.053 | 0.113 | 0.185 | 0.040 | 0.122 | 0.464 | 0.011 |

**Supplementary Table 6. Interaction effects of gestational age and DNAm CRP with brain structure; associations between DNAm CRP on global brain MRI parameters.** Standardized betas (β) and P-values are reported from regression models where DNAm CRP is regressed onto MRI measures, covarying for gestational age, sex, birthweight Z score, gestational age at scan, scanner variable (volumetric data are also corrected for head size). Additional R2 refers to the amount of variance in MRI measures accounted for DNAm CRP, beyond covariates. Bold text denotes FDR q-value <0.05.

#### Supplementary Table 7. Interaction effects of infant sex on DNAm CRP in full cohort (n=214)

| | DNAm CRP | | | | | | | DNAm_CRP x infant sex | | | | | | |
|---|---|---|---|---|---|---|---|---|---|---|---|---|---|---|
| | **b** | **Lower CI** | **Upper CI** | **p** | **r2** | **r2 (H0)** | **add r2** | **b** | **Lower CI** | **Upper CI** | **p** | **r2** | **r2 (H0)** | **add r2** |
| Cortical_grey_matter | -0.121 | -0.184 | -0.058 | 0.058 | 0.641 | 0.634 | 0.007 | 0.055 | -0.031 | 0.142 | 0.523 | 0.641 | 0.634 | 0.007 |
| Deep_grey_matter | **-0.208** | -0.271 | -0.144 | **0.001** | 0.637 | 0.618 | 0.019 | 0.127 | 0.041 | 0.214 | 0.144 | 0.637 | 0.618 | 0.019 |
| White_matter | **-0.268** | -0.348 | -0.188 | **0.001** | 0.428 | 0.396 | 0.031 | 0.166 | 0.057 | 0.275 | 0.129 | 0.428 | 0.396 | 0.031 |
| Hippocampi_and_Amygdala | **-0.206** | -0.286 | -0.125 | **0.011** | 0.421 | 0.402 | 0.018 | 0.164 | 0.054 | 0.273 | 0.137 | 0.421 | 0.402 | 0.018 |
| Cerebellum | **-0.177** | -0.244 | -0.111 | **0.008** | 0.605 | 0.591 | 0.014 | 0.126 | 0.035 | 0.216 | 0.166 | 0.605 | 0.591 | 0.014 |
| CSF | -0.047 | -0.117 | 0.024 | 0.508 | 0.554 | 0.553 | 0.001 | 0.029 | -0.067 | 0.125 | 0.764 | 0.554 | 0.553 | 0.001 |
| Ventricles | 0.051 | -0.046 | 0.148 | 0.599 | 0.153 | 0.145 | 0.008 | 0.107 | -0.026 | 0.239 | 0.421 | 0.153 | 0.145 | 0.008 |
| Brainstem | -0.071 | -0.150 | 0.008 | 0.370 | 0.440 | 0.438 | 0.002 | 0.054 | -0.054 | 0.162 | 0.616 | 0.440 | 0.438 | 0.002 |
| PSFA | **-0.166** | -0.243 | -0.088 | **0.034** | 0.458 | 0.418 | 0.041 | -0.177 | -0.283 | -0.071 | 0.097 | 0.458 | 0.418 | 0.041 |
| PSMD | **0.211** | 0.139 | 0.283 | **0.004** | 0.535 | 0.515 | 0.020 | -0.103 | -0.201 | -0.005 | 0.295 | 0.535 | 0.515 | 0.020 |
| PSAD | 0.120 | 0.034 | 0.205 | 0.163 | 0.346 | 0.339 | 0.007 | -0.056 | -0.173 | 0.060 | 0.629 | 0.346 | 0.339 | 0.007 |
| PSRD | **0.189** | 0.112 | 0.266 | **0.015** | 0.469 | 0.453 | 0.016 | -0.148 | -0.253 | -0.043 | 0.161 | 0.469 | 0.453 | 0.016 |

**Supplementary Table 7. Interaction effects of infant sex and DNAm CRP with brain structure (n= 214, term and preterm infants); associations between DNAm CRP on global brain MRI parameters.** Standardized betas (β) and P-values are reported from regression models where DNAm CRP is regressed onto MRI measures, covarying for gestational age, sex, birthweight Z score, gestational age at scan, scanner variable (volumetric data are also corrected for head size). Additional R2 refers to the amount of variance in MRI measures accounted for DNAm CRP, beyond covariates. Bold text denotes FDR q-value <0.05.

Supplementary Table 8. Interaction effects of gestational age on DNAm CRP in preterm subgroup cohort (n=127)

| | DNAm CRP | | | | | | | DNAm_CRP x gestational age | | | | | | |
|---|---|---|---|---|---|---|---|---|---|---|---|---|---|---|
| | **b** | **Lower CI** | **Upper CI** | **p** | **r2** | **r2 (H0)** | **add r2** | **b** | **Lower CI** | **Upper CI** | **p** | **r2** | **r2 (H0)** | **add r2** |
| Cortical_grey_matter | -0.134 | -0.204 | -0.065 | 0.055 | 0.563 | 0.540 | 0.023 | 0.139 | 0.063 | 0.215 | 0.071 | 0.563 | 0.540 | 0.023 |
| Deep_grey_matter | **-0.217** | -0.284 | -0.150 | **0.002** | 0.593 | 0.545 | 0.048 | **0.165** | 0.092 | 0.239 | **0.026** | 0.593 | 0.545 | 0.048 |
| White_matter | **-0.244** | -0.330 | -0.158 | **0.005** | 0.330 | 0.261 | 0.069 | **0.226** | 0.132 | 0.320 | **0.018** | 0.330 | 0.261 | 0.069 |
| Hippocampi_and_Amygdala | **-0.206** | -0.286 | -0.125 | **0.012** | 0.415 | 0.368 | 0.047 | **0.179** | 0.091 | 0.267 | **0.044** | 0.415 | 0.368 | 0.047 |
| Cerebellum | **-0.180** | -0.250 | -0.110 | **0.011** | 0.560 | 0.485 | 0.075 | **0.305** | 0.229 | 0.381 | **0.000** | 0.560 | 0.485 | 0.075 |
| CSF | -0.023 | -0.092 | 0.046 | 0.735 | 0.570 | 0.542 | 0.029 | **0.212** | 0.137 | 0.288 | **0.006** | 0.570 | 0.542 | 0.029 |
| Ventricles | 0.073 | -0.027 | 0.173 | 0.464 | 0.095 | 0.089 | 0.005 | -0.057 | -0.166 | 0.053 | 0.606 | 0.095 | 0.089 | 0.005 |
| Brainstem | -0.097 | -0.186 | -0.008 | 0.278 | 0.283 | 0.220 | 0.063 | **0.308** | 0.211 | 0.406 | **0.002** | 0.283 | 0.220 | 0.063 |
| PSFA | **-0.212** | -0.281 | -0.143 | **0.003** | 0.572 | 0.513 | 0.059 | **0.225** | 0.150 | 0.300 | **0.003** | 0.572 | 0.513 | 0.059 |
| PSMD | **0.371** | 0.283 | 0.459 | **0.000** | 0.299 | 0.166 | 0.132 | **-0.257** | -0.354 | -0.161 | **0.009** | 0.299 | 0.166 | 0.132 |
| PSAD | **0.232** | 0.147 | 0.318 | **0.007** | 0.341 | 0.263 | 0.078 | **-0.271** | -0.364 | -0.177 | **0.004** | 0.341 | 0.263 | 0.078 |
| PSRD | **0.332** | 0.236 | 0.429 | **0.001** | 0.151 | 0.054 | 0.096 | -0.182 | -0.288 | -0.076 | 0.088 | 0.151 | 0.054 | 0.096 |

**Supplementary Table 8. Interaction effects of infant sex and DNAm CRP with brain structure (n = 127, preterm infants); associations between DNAm CRP on global brain MRI parameters.** Standardized betas (β) and P-values are reported from regression models where DNAm CRP is regressed onto MRI measures, covarying for gestational age, sex, birthweight Z score, gestational age at scan, scanner variable (volumetric data are also corrected for head size). Additional R2 refers to the amount of variance in MRI measures accounted for DNAm CRP, beyond covariates. Bold text denotes FDR q-value <0.05

Supplementary Table 9. Interaction effects of gestational age on DNAm CRP in term subgroup cohort (n=85)

| | DNAm CRP | | | | | | | DNAm_CRP x gestational age | | | | | | |
|---|---|---|---|---|---|---|---|---|---|---|---|---|---|---|
| | **b** | **Lower CI** | **Upper CI** | **p** | **r2** | **r2 (H0)** | **add r2** | **b** | **Lower CI** | **Upper CI** | **p** | **r2** | **r2 (H0)** | **add r2** |
| Cortical_grey_matter | -0.033 | -0.119 | 0.054 | 0.708 | 0.496 | 0.489 | 0.007 | 0.082 | -0.006 | 0.171 | 0.353 | 0.496 | 0.489 | 0.007 |
| Deep_grey_matter | 0.008 | -0.088 | 0.103 | 0.937 | 0.389 | 0.388 | 0.001 | -0.029 | -0.126 | 0.068 | 0.765 | 0.389 | 0.388 | 0.001 |
| White_matter | -0.058 | -0.159 | 0.042 | 0.563 | 0.317 | 0.314 | 0.003 | -0.002 | -0.104 | 0.101 | 0.987 | 0.317 | 0.314 | 0.003 |
| Hippocampi_and_Amygdala | 0.092 | -0.011 | 0.195 | 0.376 | 0.281 | 0.266 | 0.016 | -0.093 | -0.198 | 0.012 | 0.379 | 0.281 | 0.266 | 0.016 |
| Cerebellum | -0.031 | -0.112 | 0.049 | 0.697 | 0.565 | 0.565 | 0.001 | -0.009 | -0.091 | 0.073 | 0.914 | 0.565 | 0.565 | 0.001 |
| CSF | -0.125 | -0.203 | -0.047 | 0.115 | 0.587 | 0.573 | 0.014 | 0.010 | -0.070 | 0.090 | 0.899 | 0.587 | 0.573 | 0.014 |
| Ventricles | -0.103 | -0.215 | 0.008 | 0.356 | 0.163 | 0.132 | 0.031 | 0.153 | 0.040 | 0.267 | 0.181 | 0.163 | 0.132 | 0.031 |
| Brainstem | -0.009 | -0.106 | 0.089 | 0.930 | 0.360 | 0.357 | 0.004 | -0.066 | -0.165 | 0.034 | 0.510 | 0.360 | 0.357 | 0.004 |
| PSFA | -0.093 | -0.180 | -0.006 | 0.287 | 0.493 | 0.475 | 0.018 | -0.121 | -0.209 | -0.032 | 0.175 | 0.493 | 0.475 | 0.018 |
| PSMD | -0.099 | -0.212 | 0.013 | 0.378 | 0.154 | 0.136 | 0.018 | 0.097 | -0.017 | 0.211 | 0.399 | 0.154 | 0.136 | 0.018 |
| PSAD | -0.097 | -0.106 | 0.188 | 0.080 | 0.203 | 0.117 | 0.086 | 0.083 | -0.027 | 0.194 | 0.454 | 0.203 | 0.117 | 0.086 |
| PSRD | -0.067 | -0.179 | 0.044 | 0.547 | 0.164 | 0.156 | 0.008 | 0.066 | -0.048 | 0.179 | 0.565 | 0.164 | 0.156 | 0.008 |

**Supplementary Table 9. Interaction effects of gestational age at birth and DNAm CRP with brain structure (n = 127, preterm infants); associations between DNAm CRP on global brain MRI parameters.** Standardized betas (β) and P-values are reported from regression models where DNAm CRP is regressed onto MRI measures, covarying for gestational age, sex, birthweight Z score, gestational age at scan, scanner variable (volumetric data are also corrected for head size). Additional R2 refers to the amount of variance in MRI measures accounted for DNAm CRP, beyond covariates. Bold text denotes FDR q-value <0.05

Supplementary table 10. Associations between global MRI metrics and DNAm CRP controlling for inflammatory exposures

| model | beta | b | Lower CI | Upper CI | pvals | r2 | additional r2 |
|---|---|---|---|---|---|---|---|
| $H_0$ (term control) | Cortical_grey_matter | -0.040 | -0.209 | 0.128 | 0.640 | 0.490 | 0.001 |
| | Deep_grey_matter | 0.010 | -0.174 | 0.195 | 0.913 | 0.388 | 0.000 |
| | White_matter | -0.058 | -0.254 | 0.137 | 0.560 | 0.317 | 0.003 |
| | Hippocampi_and_Amygdala | 0.101 | -0.100 | 0.302 | 0.328 | 0.274 | 0.009 |
| | Cerebellum | -0.031 | -0.186 | 0.125 | 0.702 | 0.565 | 0.001 |
| | CSF | -0.126 | -0.278 | 0.026 | 0.108 | 0.587 | 0.014 |
| | Ventricles | -0.118 | -0.337 | 0.101 | 0.293 | 0.144 | 0.012 |
| | Brainstem | -0.002 | -0.192 | 0.187 | 0.981 | 0.357 | 0.000 |
| | PSFA | -0.082 | -0.252 | 0.089 | 0.351 | 0.481 | 0.006 |
| | PSMD | -0.109 | -0.327 | 0.110 | 0.332 | 0.146 | 0.010 |
| | PSRD | -0.074 | -0.290 | 0.143 | 0.507 | 0.161 | 0.005 |
| | PSAD | -0.035 | -0.317 | -0.093 | 0.060 | 0.197 | 0.080 |
| $H_1$ | Cortical_grey_matter | -0.118 | -0.255 | 0.018 | 0.091 | 0.551 | 0.011 |
| | Deep_grey_matter | **-0.198** | -0.331 | -0.066 | **0.004** | 0.575 | 0.030 |
| | White_matter | **-0.218** | -0.389 | -0.048 | **0.013** | 0.298 | 0.037 |
| | Hippocampi_and_Amygdala | **-0.185** | -0.343 | -0.027 | **0.024** | 0.395 | 0.027 |
| | Cerebellum | -0.145 | -0.289 | -0.001 | 0.050 | 0.501 | 0.016 |
| | CSF | 0.001 | -0.137 | 0.139 | 0.989 | 0.542 | 0.000 |
| | Ventricles | 0.067 | -0.127 | 0.261 | 0.500 | 0.093 | 0.003 |
| | Brainstem | -0.062 | -0.241 | 0.118 | 0.502 | 0.222 | 0.003 |
| | PSFA | **-0.186** | -0.324 | -0.048 | **0.009** | 0.540 | 0.027 |
| | PSMD | **0.341** | 0.166 | 0.517 | **0.000** | 0.256 | 0.090 |
| | PSRD | **0.312** | 0.122 | 0.501 | **0.002** | 0.130 | 0.075 |
| | PSAD | **0.201** | 0.030 | 0.372 | **0.023** | 0.294 | 0.031 |
| $H_2$ (fully adjusted model) | Cortical_grey_matter | -0.141 | -0.298 | 0.016 | 0.081 | 0.589 | 0.011 |
| | Deep_grey_matter | **-0.209** | -0.362 | -0.057 | **0.008** | 0.613 | 0.025 |
| | White_matter | **-0.304** | -0.502 | -0.107 | **0.003** | 0.351 | 0.052 |
| | Hippocampi_and_Amygdala | -0.136 | -0.324 | 0.052 | 0.160 | 0.412 | 0.010 |
| | Cerebellum | **-0.204** | -0.362 | -0.046 | **0.013** | 0.585 | 0.024 |
| | CSF | 0.022 | -0.136 | 0.180 | 0.785 | 0.583 | 0.000 |
| | Ventricles | -0.030 | -0.252 | 0.192 | 0.792 | 0.182 | 0.001 |

| | | | | | | | |
|---|---|---|---|---|---|---|---|
| | Brainstem | -0.096 | -0.304 | 0.112 | 0.369 | 0.279 | 0.005 |
| | PSFA | **-0.215** | -0.375 | -0.055 | **0.009** | 0.577 | 0.026 |
| | PSMD | **0.206** | 0.009 | 0.403 | **0.042** | 0.357 | 0.024 |
| | PSRD | 0.175 | -0.041 | 0.392 | 0.115 | 0.222 | 0.017 |
| | PSAD | 0.093 | -0.095 | 0.280 | 0.336 | 0.414 | 0.005 |
| $H_3$ (maternal smoking in pregnancy) | Cortical_grey_matter | -0.118 | -0.255 | 0.018 | 0.092 | 0.551 | 0.011 |
| | Deep_grey_matter | **-0.199** | -0.331 | -0.067 | **0.004** | 0.581 | 0.031 |
| | White_matter | **-0.218** | -0.390 | -0.047 | **0.014** | 0.298 | 0.037 |
| | Hippocampi_and_Amygdala | **-0.186** | -0.344 | -0.027 | **0.023** | 0.397 | 0.027 |
| | Cerebellum | -0.145 | -0.289 | -0.001 | 0.051 | 0.501 | 0.016 |
| | CSF | 0.001 | -0.137 | 0.139 | 0.994 | 0.543 | 0.000 |
| | Ventricles | 0.067 | -0.128 | 0.261 | 0.502 | 0.093 | 0.003 |
| | Brainstem | -0.062 | -0.242 | 0.117 | 0.498 | 0.227 | 0.003 |
| | PSFA | **-0.187** | -0.325 | -0.048 | **0.009** | 0.541 | 0.027 |
| | PSMD | **0.343** | 0.168 | 0.517 | **0.000** | 0.273 | 0.091 |
| | PSRD | **0.313** | 0.124 | 0.502 | **0.002** | 0.145 | 0.076 |
| | PSAD | **0.202** | 0.032 | 0.373 | **0.022** | 0.303 | 0.032 |
| $H_4$ (preeclampsia) | Cortical_grey_matter | -0.116 | -0.252 | 0.020 | 0.097 | 0.557 | 0.010 |
| | Deep_grey_matter | **-0.197** | -0.329 | -0.064 | **0.004** | 0.579 | 0.030 |
| | White_matter | **-0.216** | -0.386 | -0.046 | **0.014** | 0.304 | 0.036 |
| | Hippocampi_and_Amygdala | **-0.184** | -0.343 | -0.025 | **0.025** | 0.396 | 0.026 |
| | Cerebellum | -0.143 | -0.287 | 0.001 | 0.053 | 0.505 | 0.016 |
| | CSF | 0.002 | -0.136 | 0.140 | 0.972 | 0.544 | 0.000 |
| | Ventricles | 0.064 | -0.130 | 0.257 | 0.520 | 0.103 | 0.003 |
| | Brainstem | -0.057 | -0.235 | 0.121 | 0.530 | 0.244 | 0.003 |
| | PSFA | **-0.183** | -0.320 | -0.046 | **0.010** | 0.550 | 0.026 |
| | PSMD | **0.338** | 0.163 | 0.513 | **0.000** | 0.268 | 0.088 |
| | PSRD | **0.309** | 0.119 | 0.499 | **0.002** | 0.137 | 0.074 |
| | PSAD | **0.198** | 0.028 | 0.368 | **0.024** | 0.307 | 0.030 |
| $H_5$ (HCA) | Cortical_grey_matter | -0.120 | -0.257 | 0.017 | 0.089 | 0.552 | 0.011 |
| | Deep_grey_matter | **-0.196** | -0.329 | -0.063 | **0.005** | 0.577 | 0.030 |
| | White_matter | **-0.220** | -0.391 | -0.049 | **0.013** | 0.299 | 0.037 |
| | Hippocampi_and_Amygdala | **-0.181** | -0.339 | -0.023 | **0.027** | 0.400 | 0.025 |
| | Cerebellum | -0.142 | -0.286 | 0.002 | 0.056 | 0.504 | 0.016 |
| | CSF | 0.009 | -0.127 | 0.144 | 0.901 | 0.560 | 0.000 |

| | | | | | | | |
|---|---|---|---|---|---|---|---|
| | Ventricles | 0.071 | -0.123 | 0.265 | 0.475 | 0.098 | 0.004 |
| | Brainstem | -0.053 | -0.230 | 0.125 | 0.561 | 0.247 | 0.002 |
| | PSFA | **-0.186** | -0.325 | -0.048 | **0.010** | 0.540 | 0.027 |
| | PSMD | **0.341** | 0.164 | 0.517 | **0.000** | 0.257 | 0.090 |
| | PSRD | **0.311** | 0.120 | 0.502 | **0.002** | 0.130 | 0.075 |
| | PSAD | **0.203** | 0.032 | 0.375 | **0.022** | 0.295 | 0.032 |
| | Cortical_grey_matter | -0.119 | -0.255 | 0.018 | 0.091 | 0.555 | 0.011 |
| | Deep_grey_matter | **-0.198** | -0.330 | -0.067 | **0.004** | 0.585 | 0.030 |
| | White_matter | **-0.218** | -0.389 | -0.048 | **0.014** | 0.302 | 0.037 |
| | Hippocampi_and_Amygdala | **-0.185** | -0.344 | -0.027 | **0.024** | 0.397 | 0.027 |
| | Cerebellum | **-0.145** | -0.283 | -0.007 | **0.042** | 0.543 | 0.016 |
| $H_6$ (ROP) | CSF | 0.001 | -0.135 | 0.137 | 0.990 | 0.558 | 0.000 |
| | Ventricles | 0.067 | -0.128 | 0.261 | 0.502 | 0.093 | 0.003 |
| | Brainstem | -0.062 | -0.241 | 0.117 | 0.500 | 0.233 | 0.003 |
| | PSFA | **-0.186** | -0.322 | -0.051 | **0.008** | 0.558 | 0.027 |
| | PSMD | **0.342** | 0.170 | 0.513 | **0.000** | 0.296 | 0.090 |
| | PSRD | **0.312** | 0.124 | 0.500 | **0.001** | 0.153 | 0.075 |
| | PSAD | **0.201** | 0.036 | 0.367 | **0.019** | 0.342 | 0.031 |
| | Cortical_grey_matter | -0.098 | -0.240 | 0.044 | 0.177 | 0.555 | 0.007 |
| | Deep_grey_matter | **-0.218** | -0.356 | -0.080 | **0.002** | 0.579 | 0.034 |
| | White_matter | **-0.245** | -0.423 | -0.068 | **0.008** | 0.305 | 0.043 |
| | Hippocampi_and_Amygdala | **-0.185** | -0.351 | -0.019 | **0.030** | 0.395 | 0.024 |
| | Cerebellum | -0.111 | -0.260 | 0.038 | 0.146 | 0.511 | 0.009 |
| $H_7$ (NEC) | CSF | 0.035 | -0.108 | 0.177 | 0.635 | 0.552 | 0.001 |
| | Ventricles | 0.098 | -0.104 | 0.299 | 0.344 | 0.101 | 0.007 |
| | Brainstem | -0.051 | -0.239 | 0.136 | 0.593 | 0.223 | 0.002 |
| | PSFA | **-0.199** | -0.343 | -0.054 | **0.008** | 0.541 | 0.028 |
| | PSMD | **0.292** | 0.112 | 0.473 | **0.002** | 0.278 | 0.061 |
| | PSRD | **0.252** | 0.057 | 0.447 | **0.012** | 0.161 | 0.045 |
| | PSAD | **0.201** | 0.022 | 0.380 | **0.029** | 0.294 | 0.029 |
| | Cortical_grey_matter | **-0.180** | -0.324 | -0.036 | **0.015** | 0.571 | 0.030 |
| | Deep_grey_matter | **-0.243** | -0.384 | -0.102 | **0.001** | 0.586 | 0.041 |
| $H_8$ (Sepsis) | White_matter | **-0.304** | -0.482 | -0.125 | **0.001** | 0.335 | 0.074 |
| | Hippocampi_and_Amygdala | **-0.183** | -0.353 | -0.012 | **0.038** | 0.395 | 0.027 |
| | Cerebellum | **-0.223** | -0.373 | -0.073 | **0.004** | 0.532 | 0.047 |

| | | | | | | | |
|---|---|---|---|---|---|---|---|
| | CSF | -0.032 | -0.179 | 0.116 | 0.675 | 0.547 | 0.005 |
| | Ventricles | -0.024 | -0.228 | 0.179 | 0.816 | 0.135 | 0.046 |
| | Brainstem | -0.119 | -0.310 | 0.072 | 0.225 | 0.239 | 0.020 |
| | PSFA | **-0.218** | -0.366 | -0.071 | **0.004** | 0.545 | 0.032 |
| | PSMD | **0.270** | 0.084 | 0.455 | **0.005** | 0.283 | 0.116 |
| | PSRD | **0.225** | 0.026 | 0.425 | **0.029** | 0.168 | 0.113 |
| | PSAD | 0.172 | -0.011 | 0.356 | 0.068 | 0.298 | 0.036 |
| $H_9$ (BPD) | Cortical_grey_matter | -0.093 | -0.239 | 0.053 | 0.213 | 0.555 | 0.006 |
| | Deep_grey_matter | **-0.154** | -0.295 | -0.013 | **0.034** | 0.586 | 0.016 |
| | White_matter | **-0.199** | -0.382 | -0.016 | **0.035** | 0.300 | 0.027 |
| | Hippocampi_and_Amygdala | -0.147 | -0.317 | 0.022 | 0.090 | 0.402 | 0.015 |
| | Cerebellum | **-0.160** | -0.315 | -0.006 | **0.044** | 0.502 | 0.017 |
| | CSF | 0.011 | -0.137 | 0.160 | 0.880 | 0.542 | 0.000 |
| | Ventricles | 0.031 | -0.176 | 0.239 | 0.768 | 0.099 | 0.001 |
| | Brainstem | -0.067 | -0.260 | 0.126 | 0.495 | 0.223 | 0.003 |
| | PSFA | **-0.175** | -0.323 | -0.026 | **0.023** | 0.540 | 0.021 |
| | PSMD | **0.303** | 0.116 | 0.491 | **0.002** | 0.264 | 0.062 |
| | PSRD | **0.292** | 0.088 | 0.496 | **0.006** | 0.132 | 0.058 |
| | PSAD | 0.115 | -0.064 | 0.294 | 0.210 | 0.333 | 0.009 |

**Supplementary table 10. Associations between DNAm CRP on global brain MRI parameters, adjusting for inflammatory exposures.** Standardized betas (β) and P-values are reported from regression models where DNAm CRP is regressed onto MRI measures, additional R2 refers to the amount of variance in MRI measures accounted for DNAm CRP, beyond covariates. Bold text denotes FDR q-value <0.05. Models are as follows:

Model $H_0$ = MRI metric ~ DNAm CRP + GA birth, infant sex, birthweight, GA scan, scanner variable (term subgroup, n = 87)

Model $H_1$ = MRI metric ~ DNAm CRP + GA birth, infant sex, birthweight, GA scan, scanner variable (preterm subgroup, n = 127)

Model $H_3$ = MRI metric ~ DNAm CRP + GA birth, infant sex, birthweight, GA scan, scanner variable + maternal smoking in pregnancy (preterm subgroup, n = 127)

Model $H_4$ = MRI metric ~ DNAm CRP + GA birth, infant sex, birthweight, GA scan, scanner variable + preeclampsia (preterm subgroup, n = 127)

Model $H_5$ = MRI metric ~ DNAm CRP + GA birth, infant sex, birthweight, GA scan, scanner variable + HCA (preterm subgroup, n = 127)

Model $H_6$ = MRI metric ~ DNAm CRP + GA birth, infant sex, birthweight, GA scan, scanner variable + ROP (preterm subgroup, n = 127)

Model $H_7$ = MRI metric ~ DNAm CRP + GA birth, infant sex, birthweight, GA scan, scanner variable + NEC (preterm subgroup, n = 127)

Model $H_8$ = MRI metric ~ DNAm CRP + GA birth, infant sex, birthweight, GA scan, scanner variable + Sepsis (preterm subgroup, n = 127)

Model $H_9$ = MRI metric ~ DNAm CRP + GA birth, infant sex, birthweight, GA scan, scanner variable + BPD (preterm subgroup, n = 127)

Supplementary table 11. Associations between individual DTI tract metrics and DNAm CRP controlling for inflammatory exposures

| model | Tract | FA | | | | | | MD | | | | | |
|---|---|---|---|---|---|---|---|---|---|---|---|---|---|
| | | beta | lower CI | upper CI | pvals | r2 | add_r2 | beta | lower CI | upper CI | pvals | r2 | add_r2 |
| $H_1$ | AF left | **-0.433** | -0.687 | -0.179 | **1.45E-03** | 0.223 | 0.150 | **0.310** | 0.051 | 0.570 | **0.023** | 0.188 | 0.077 |
| | AF right | **-0.434** | -0.677 | -0.190 | **9.32E-04** | 0.285 | 0.150 | **0.331** | 0.063 | 0.598 | **0.019** | 0.137 | 0.087 |
| | ATR left | **-0.380** | -0.631 | -0.129 | **4.37E-03** | 0.242 | 0.115 | **0.392** | 0.164 | 0.620 | **0.001** | 0.375 | 0.122 |
| | ATR right | -0.261 | -0.517 | -0.004 | 0.051 | 0.207 | 0.054 | 0.249 | -0.004 | 0.502 | 0.058 | 0.229 | 0.050 |
| | CC genu | **-0.381** | -0.596 | -0.166 | **9.93E-04** | 0.443 | 0.116 | **0.472** | 0.224 | 0.720 | **0.000** | 0.260 | 0.178 |
| | CC splenium | -0.119 | -0.371 | 0.133 | 0.357 | 0.234 | 0.011 | 0.221 | -0.040 | 0.482 | 0.103 | 0.178 | 0.039 |
| | CCG left | **-0.325** | -0.588 | -0.062 | **0.019** | 0.167 | 0.084 | **0.332** | 0.082 | 0.582 | **0.012** | 0.248 | 0.088 |
| | CCG right | **-0.455** | -0.718 | -0.191 | **1.28E-03** | 0.165 | 0.165 | **0.393** | 0.131 | 0.654 | **0.005** | 0.175 | 0.123 |
| | CST left | **-0.411** | -0.638 | -0.184 | **7.91E-04** | 0.377 | 0.135 | **0.408** | 0.168 | 0.648 | **0.002** | 0.306 | 0.133 |
| | CST right | **-0.549** | -0.773 | -0.326 | **1.06E-05** | 0.400 | 0.241 | **0.469** | 0.245 | 0.693 | **0.000** | 0.395 | 0.176 |
| | IFOF left | **-0.391** | -0.629 | -0.153 | **0.002** | 0.320 | 0.122 | **0.337** | 0.100 | 0.574 | **0.007** | 0.325 | 0.091 |
| | IFOF right | **-0.414** | -0.642 | -0.185 | **7.90E-04** | 0.369 | 0.136 | **0.283** | 0.028 | 0.538 | **0.034** | 0.216 | 0.064 |
| | ILF left | **-0.285** | -0.515 | -0.055 | **0.018** | 0.361 | 0.065 | **0.282** | 0.036 | 0.529 | **0.029** | 0.266 | 0.064 |
| | ILF right | **-0.337** | -0.581 | -0.093 | **8.92E-03** | 0.283 | 0.091 | 0.237 | -0.030 | 0.504 | 0.087 | 0.141 | 0.045 |
| | UNC left | **-0.421** | -0.659 | -0.182 | **1.02E-03** | 0.315 | 0.141 | **0.355** | 0.137 | 0.573 | **0.002** | 0.426 | 0.100 |
| | UNC right | **-0.384** | -0.625 | -0.143 | **2.76E-03** | 0.302 | 0.118 | **0.321** | 0.079 | 0.563 | **0.012** | 0.294 | 0.082 |
| $H_2$ | AF left | -0.251 | -0.564 | 0.062 | 0.123 | 0.310 | 0.033 | 0.205 | -0.120 | 0.530 | 0.221 | 0.257 | 0.022 |
| | AF right | **-0.434** | -0.733 | -0.136 | **0.006** | 0.373 | 0.100 | 0.272 | -0.059 | 0.602 | 0.113 | 0.232 | 0.039 |
| | ATR left | -0.208 | -0.511 | 0.095 | 0.184 | 0.356 | 0.023 | **0.375** | 0.100 | 0.651 | **0.010** | 0.466 | 0.075 |
| | ATR right | -0.234 | -0.546 | 0.079 | 0.149 | 0.315 | 0.029 | 0.135 | -0.159 | 0.428 | 0.373 | 0.395 | 0.010 |
| | CC genu | -0.185 | -0.438 | 0.069 | 0.160 | 0.548 | 0.018 | **0.367** | 0.084 | 0.649 | **0.014** | 0.440 | 0.071 |
| | CC splenium | -0.148 | -0.453 | 0.158 | 0.348 | 0.344 | 0.012 | 0.087 | -0.218 | 0.393 | 0.577 | 0.345 | 0.004 |
| | CCG left | -0.184 | -0.505 | 0.137 | 0.267 | 0.276 | 0.018 | **0.376** | 0.060 | 0.693 | **0.024** | 0.297 | 0.075 |
| | CCG right | **-0.406** | -0.735 | -0.078 | **0.019** | 0.241 | 0.087 | **0.464** | 0.146 | 0.782 | **0.006** | 0.289 | 0.114 |
| | CST left | **-0.344** | -0.623 | -0.066 | **0.019** | 0.455 | 0.063 | **0.305** | 0.028 | 0.582 | **0.035** | 0.462 | 0.049 |
| | CST right | **-0.540** | -0.822 | -0.257 | **4.65E-04** | 0.438 | 0.154 | **0.368** | 0.102 | 0.634 | **0.009** | 0.504 | 0.072 |
| | IFOF left | -0.189 | -0.434 | 0.056 | 0.137 | 0.577 | 0.019 | 0.165 | -0.093 | 0.422 | 0.216 | 0.534 | 0.014 |

| | | | | | | | | | | | | |
|---|---|---|---|---|---|---|---|---|---|---|---|---|
| **IFOF right** | **-0.313** | -0.570 | -0.056 | **0.021** | 0.535 | 0.052 | **0.294** | -0.005 | 0.592 | **0.059** | 0.375 | 0.046 |
| **ILF left** | -0.141 | -0.406 | 0.124 | 0.302 | 0.507 | 0.010 | 0.134 | -0.132 | 0.399 | 0.329 | 0.504 | 0.009 |
| **ILF right** | -0.269 | -0.557 | 0.018 | 0.072 | 0.419 | 0.038 | 0.255 | -0.046 | 0.556 | 0.104 | 0.363 | 0.034 |
| **UNC left** | -0.270 | -0.537 | -0.002 | 0.053 | 0.498 | 0.039 | **0.370** | 0.105 | 0.635 | **0.009** | 0.505 | 0.072 |
| **UNC right** | -0.192 | -0.455 | 0.071 | 0.158 | 0.513 | 0.020 | **0.321** | 0.038 | 0.605 | **0.031** | 0.436 | 0.055 |

**Supplementary Table 11. Associations between DNAm CRP and the dMRI metrics of FA and MD in each tract.** The β coefficients are in the units of standard deviations. Reported p-values are adjusted for false discovery rate (FDR).
